# Estimating the worst-case scenario for malaria parasite rate in sub-Saharan Africa

**DOI:** 10.1101/2025.05.16.25327745

**Authors:** Kaustubh Chakradeo, Alexandros Katsiferis, Neil Scheidwasser, Iwona Hawryluk, Katherine E Battle, Swapnil Mishra, David L Smith, Seth Flaxman, David Duchene, Samir Bhatt

**Affiliations:** Section of Health Data Science and AI, University of Copenhagen, Copenhagen, Denmark; Department of Infectious Disease Epidemiology, Imperial College London, London, United Kingdom; Institute for Disease Modelling, Gates Foundation, Seattle, Washington, United States of America; Saw Swee Hock School of Public Health and Institute of Data Science, National University of Singapore, Singapore, Singapore; Department of Health Metrics Science, University of Washington, Seattle, Washington, United States of America; Department of Computer Science, University of Oxford, Oxford, United Kingdom; MRC Centre for Global Infectious Disease Analysis, Imperial College London, London, United Kingdom

**Keywords:** Malaria Baseline, Sub-Saharan Africa, Contrastive Deep Learning, Satellite Imagery

## Abstract

Malaria remains a leading cause of morbidity and mortality worldwide, with sub-Saharan Africa bearing the highest burden. Stalled progress under an inadequate budget and the expectation that funding would be further constrained [1, 2] call for an evaluation of a worst-case scenario to appreciate malaria transmission potential in the absence of interventions. We examine a scenario in which all funding for interventions ceases for several years, allowing population immunity to adjust to a new equilibrium amid a surge in transmission, while economic development, climate, and demographics remain static, which we define as the transmission niche or “baseline” prevalence. The baseline proposed here is a set of stratified *PfPr*_2-10_ observations to select only samples with low intervention histories, preserving diverse epidemiological context. We further developed a bespoke contrastive deep learning architecture applied to 30-meter resolution images from Landsat 8 satellites, generating a detailed vector covariate set based on observable land status, which helps produce state-of-the-art malaria estimates, an improvement over all previous covariate-based models. These features were integrated into a Bayesian MCMC model with regularisation to estimate baseline *PfPr*_2-10_ and ***R*_0_** to generate a map of malaria incidence over all of Sub-Saharan Africa. Using population data from the World Malaria Report 2024, our model estimates 422 (275–582) million cases across sub-Saharan Africa for this worst-case baseline scenario, reflecting a 131% increase compared to previous business-as-usual baseline scenario estimates [3]. Our analyses highlight the effect of long-term benefits of two decades of investments for malaria control and the critical need for sustained intervention efforts and informed policy-making to mitigate potential resurgences in malaria transmission.

## Main

Malaria remains one of the deadliest infectious diseases globally, causing over 250M cases and 550K deaths in 2023 [4]. Malaria transmission and prevalence in a population, including its intensity and seasonality, is influenced by a range of complex, interdependent factors. These include: weather conditions [5, 6], mosquito ecology [7], biogeography [8], malaria control efforts [3], economic development initiatives [9–13], and historical events [14]. During the 20^th^ and 21^st^ centuries, malaria prevalence has fluctuated dramatically across space and time, but there has been a broad trend of transmission and mortality decline coupled with a substantially reduced geographical range [5, 15]. These changes have coincided with significant developments in public health and have been closely intertwined with economic progress and efforts aimed at malaria control. Examples include the Abuja declaration [16], the establishment of the Global Malaria Action Plan in 2008 (a first comprehensive strategy for the implementation of malaria elimination programs) [1, 2], the establishment and enhancement of health information systems [17], the utilisation of medical interventions for malaria control [18], the emergence of trends linked to the evolution of drug resistance [19], and the evolution of health infrastructure [20, 21]. Despite these efforts at intervention coverage aimed at controlling malaria, malaria outbreaks remain common [22–25], and are driven by the evolution of insecticide resistance or the cessation of malaria control efforts.

Against this backdrop of changing malaria burden, it is likely that the continued fight against malaria is at a tipping point [26], and progress in Sub-Saharan Africa (SSA) in particular has stalled [27]. Achieving the targets of WHO’s Global Technical Strategy for Malaria 2016–2030 [28] will require innovative scientific and programmatic decisions at a time when the funding landscape is increasingly constrained. A crucial scientific endeavour to calibrate and anchor global resolve is to estimate a reasonable worst-case scenario of malaria prevalence in SSA should all funding for interventions in SSA cease for several years, population immunity levels reached a new equilibrium in the subsequent surge in malaria transmission, but the economic development, climate, population, and other aspects remained static. Following ecological parlance, we refer to this scenario as the transmission niche or “baseline” prevalence. In other words, what would the expected malaria prevalence be in SSA now, in the absence of interventions?

Previous approaches for mapping malaria have focused on creating contemporary maps or time series [3, 5, 15, 22, 29] using parasite rates (prevalence) collected in household surveys and/or routine surveillance data capturing malaria cases diagnosed in healthcare facilities. When parasite rate data are used as the response variable, adjustments are made to standardise the input data for malaria diagnostic test and the age structure of the sample population, with the latter often converted to the 2-10 age range (that is, a common measure of *Plasmodium falciparum* transmission intensity, *PfPr* _2-10_) [30, 31]. Previous studies on malaria mapping employed Gaussian process regression for spatial and temporal mapping [32], with a suite of temporally varying and gap-filled (for cloud cover and missing data) environmental covariates [33]. These covariates can be used as a linear basis [34], transformed [35] or stacked/super-learned [36]. Additionally, none of the previous studies were designed to formally evaluate temporal changes in malaria because of interventions [3]. From these previous models, it is possible to project backwards in time to estimate the transmission levels at some historical period for which interpolation is valid given the data [3]. This approach has limitations: First, because any given country typically had a different historical distribution of interventions, it is challenging to estimate a baseline level of *PfPr* _2-10_ for all of SSA. This is mainly due to the temporal heterogeneity at the beginning of widespread intervention use, as in the cases of insecticide-treated bed nets (ITN), indoor residual spraying (IRS), or the switch of frontline antimalarial drugs to artemisinin combination therapy (ACT). Second, while Gaussian processes are excellent at interpolation, the need to subset data due to the temporal heterogeneity in interventions means that *PfPr* _2-10_ will be sparse in both space and time, with substantially smaller datasets than those used in standard large mapping efforts [3, 15, 37]. In addition, such data sparsity might pose a risk of sampling bias, which could lead to a biased estimate in the space-time random field of the Gaussian process and, in turn, the estimated map. Third, if a Gaussian process is not used, then the accuracy and generalizability of prediction from mapping through malaria-related covariates alone is a limitation [35]. This is due to both the environmental and land use information contained in the covariates and their coarse resolution relative to heterogeneous landscape features that influence vector habitat. Furthermore, the choice of covariates is often arbitrary and depends on availability or prepossessed products (e.g., [38]).

Here, we attempt to overcome limitations in previous efforts to define baseline *PfPr* prevalence by first stratifying a full database of *PfPr* _2-10_ observations to select only those with a low intervention history, such as areas with minimal or no distribution of insecticide-treated nets, or access to antimalarial treatments. This step is taken to retain parasite rate samples from a variety of epidemiological contexts while avoiding the confounding effects of interventions, ensuring that the selected data more accurately reflects the natural transmission dynamics of malaria. To create more meaningful covariates, we develop a bespoke deep learning architecture, which we trained using a contrastive learning paradigm [39–42] over 30-meter resolution images from Landsat 8 [43] to learn meaningful covariates from raw satellite imagery. This approach offers several advantages: it provides state-of-the-art performance, an improvement over previous covariate-based results; it models environmental covariates more effectively; and it eliminates the issue of missing covariates, which is common with surveys or handcrafted covariates, as these covariates are derived directly from transformed satellite images. In contrast to previous approaches that define a single covariate value for a 5km or 10km square, we preprocess 30-meter resolution images to yield a much expanded covariate set that is not arbitrarily defined but based on observable land status within remotely sensed imagery. Using the deep learning features generated from these images, we utilise Bayesian MCMC sampling [44, 45] with Bayesian regularisation to estimate a map of baseline *PfPr* _2-10_ and *R*_0_. The complete steps for the estimation of baseline malaria incidence are described in Figure 1.

**Fig. 1:**
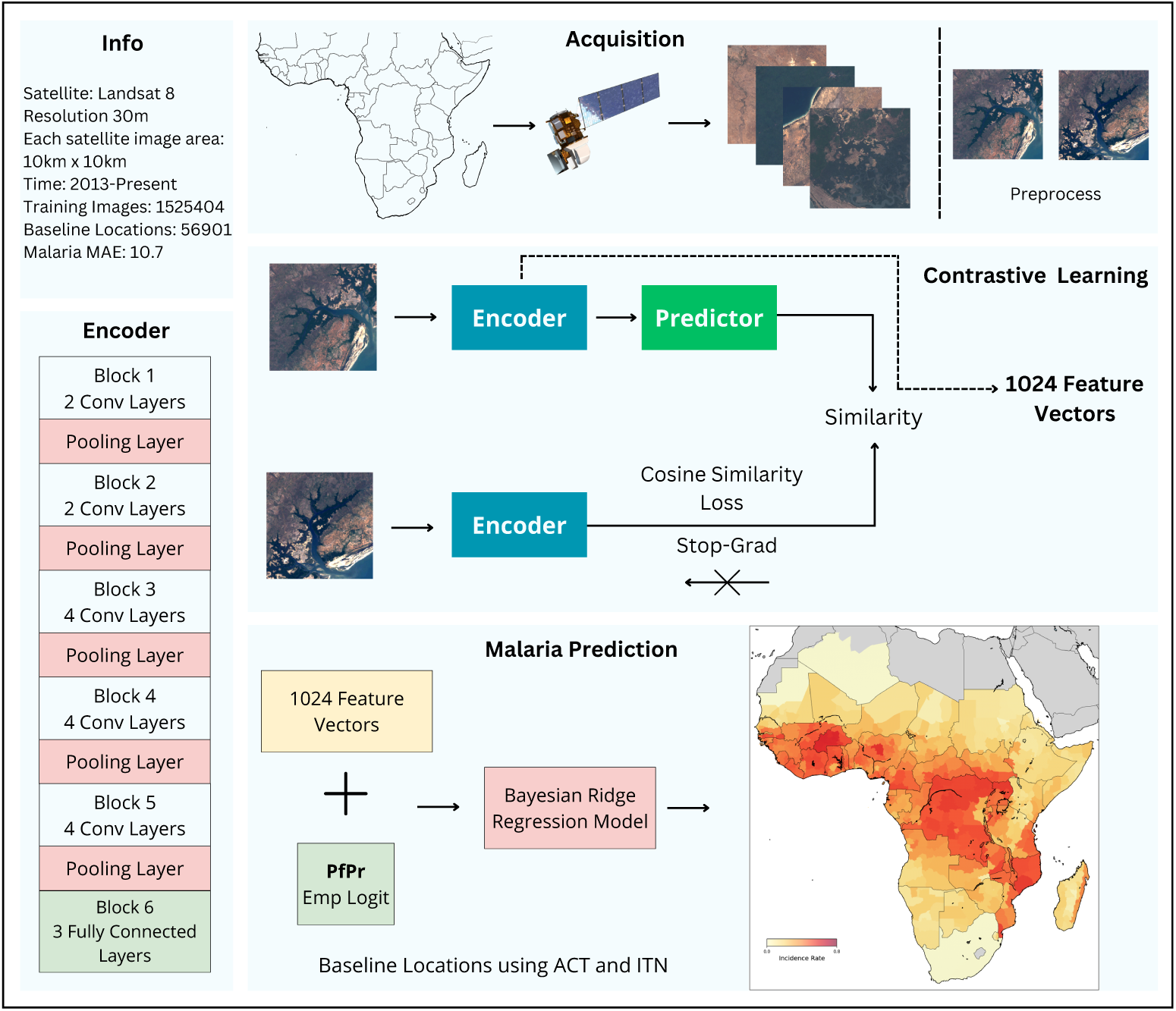
Estimates of the baseline parasite rate of Malaria in SSA. We first acquired 30m resolution Landsat 8 images from Google Earth Engine from the beginning of 2013 until 2024. Each image captured a 100km^2^ land area. Subsequently, we train a convolutional neural network (CNN) architecture using a contrastive deep learning paradigm [39]. In the contrastive paradigm, for each image, we perform data augmentation to create two augmented views of the input, which are then fed to an encoder network. During training, a predictor is trained to maximise the similarity between the two encoder outputs. During inference, the images are simply fed to the encoder to extract a 1024-dimensional embedding. Using locations in sub-Saharan Africa with a coverage of less than 10% insecticide-treated bed-nets and less than 50% Artemisinin-based combination treatments for malaria, and selecting points before the establishment of the Global Malaria Action Plan in 2008 [1, 2], providing a baseline of locations before widespread reporting of malaria interventions were in place. The feature vectors from the contrastive learning model were then used to estimate the prevalence of malaria for these baseline points. Before predictions, the malaria prevalence rate was converted to an empirical logit scale. Finally, a malaria incidence map for sub-Saharan Africa was produced using the estimated malaria baseline and populations from 2024.

## Results

First, we generated feature vectors for all of SSA using a bespoke convolutional neural network trained using contrastive learning [39]. These vector embeddings were designed to generically identify environmental and land-use features such as water bodies, forests, vegetation, elevation, deserts, urban settlements and road networks. For example, regressing our features against known data on enhanced vegetation indices from the MODIS satellite showed excellent correspondence, highlighting how the contrastive model captures a rich range of features generically with a mean absolute error of 0.03 and correlation of 0.93. Each 10×10km square across SSA, excluding several Northern African countries which are malaria-free, was associated with a satellite image encoded as a 1024-dimensional vector that generically represents the features present in that image.

From all the points in SSA, we used a threshold to construct our baseline. These baseline points consisted of retrospective locations which had low anti-malarial drug usage, and low insectide treated nets/indoor residual spraying adoption (supplementary Figure 11). Using a Bayesian regression model, malaria prevalence rates were estimated for the baseline points with the contrastive features as the covariate variables, and *PfPr* _2-10_ prevalence as the dichotomous binomial response variable. Using this predictive regression model, we were able to estimate *PfPr* _2-10_ prevalence rates with uncertainty for all of SSA. These prevalence rates were converted to population-weighted incidence rates using the WorldPop population raster from 2023 [46]. Finally, from these incidence rates, Admin 0 and Admin 1 level estimates for incidence rates were generated via aggregation using conditional sampling and propagation of uncertainty. These rates represent an estimate of baseline malaria (Figure 2); these are the estimated incidence rates if malaria interventions were stopped for a long period of time and political, environmental and demographic changes did not occur. Figure 2 shows the full malaria incidence map estimated using the generated baseline malaria points from the contrastive deep learning model. The mean absolute error for SSA was 10.7%, which is a large improvement on previous covariate-based estimates[35] and broadly comparable with a full Gaussian process regression model.

**Fig. 2:**
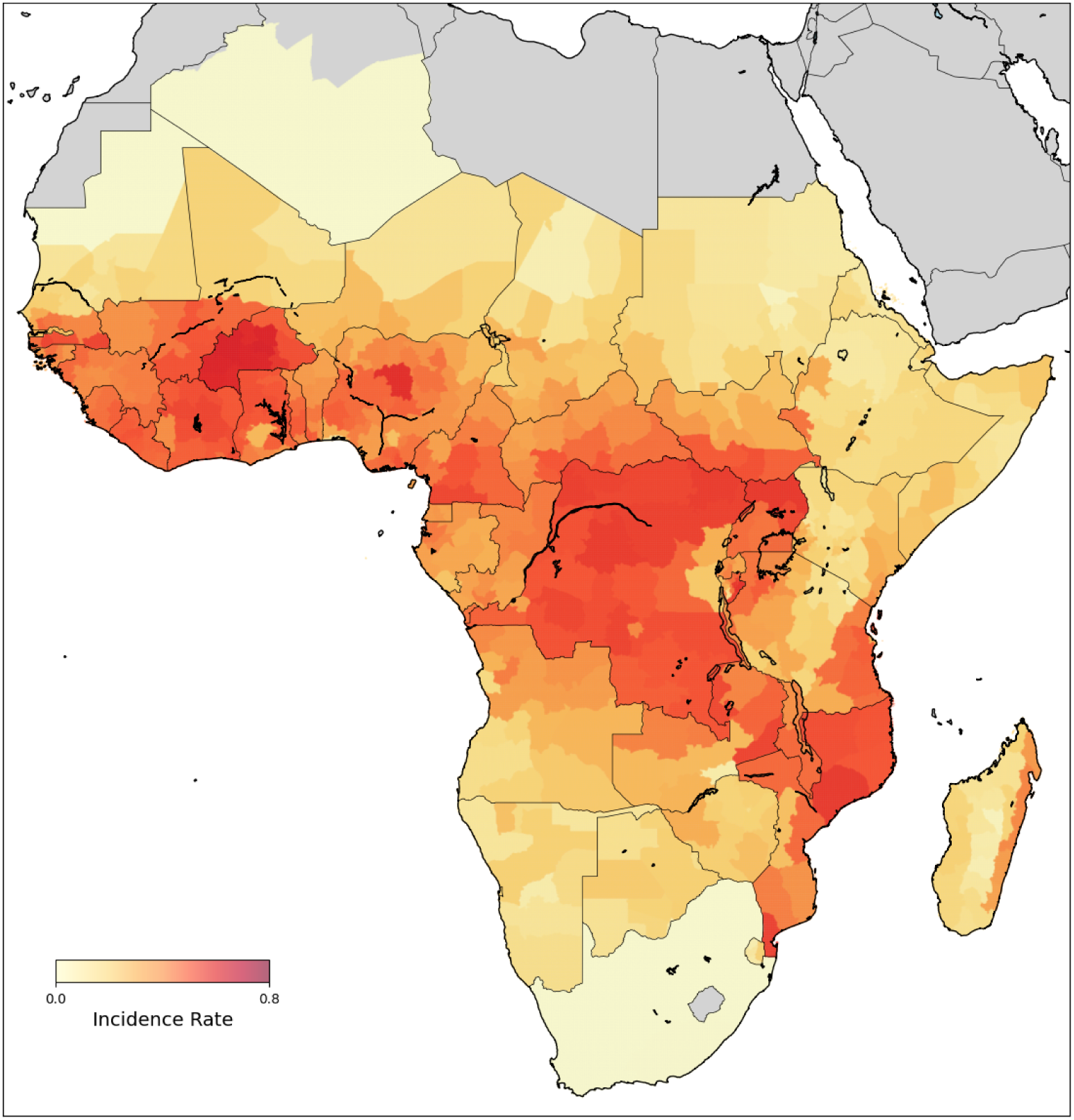
Estimated Baseline *P. falciparum* Incidence Rate, according to WMR 2024 population numbers.

### Estimating Baseline Malaria Rate

By using the baseline, we first estimate the mean malaria prevalence based on 200 realisations from the Bayesian model with features from the deep learning model as covariates. We then convert this prevalence to incidence rates [46] (see Methods) and then plot the estimated incidence rates of the proportion of the population infected based on population denominator from WMR 2024 across SSA if interventions were halted (Figure 2). The estimated malaria incidence is highest in West Africa and Central Africa, as well as around Lake Victoria, with Nigeria, Burkina Faso and Ghana, the Democratic Republic of Congo, Uganda, Kenya and Tanzania particularly affected. These regions are mainly densely populated rural and peri-urban regions and exhibit the highest malaria rates. If interventions were stopped, population densities would play a key role in the spread of malaria, and this map exhibits the same.

Since the year 2000, there has been a sustained effort of malaria interventions across SSA. However, in recent years and especially after 2015, the funding for malaria has plateaued and coverage of vector control interventions has declined [27, 47]. The model shows that malaria incidence rates will increase if funding and intervention keep sliding down. Despite the best efforts of interventions for the last 2 decades, our data show that malaria incidence *will* see a widespread increase regardless of the current incidence rates. In SSA, we estimate there will be 422.3 (95% UI 275.2 - 582.5) million malaria cases if all interventions ceased for several years (from 1 year for some countries to 15 years for some, based on the year of intervention coverage (supplementary Figure 11)), assuming development, climate and population remain stable (Table 1). As compared to the latest numbers from the World Malaria Report 2024 (WMR 2024), in every single country, there would be an increase in malaria cases without interventions, with Nigeria (79.26 million), DRC (26.62 million) and Tanzania (23.48 million) showing the highest increase in malaria cases (Figure 3). Even countries with low numbers in 2024 would see an explosive percentage rise in the estimated cases if interventions ceased. Even though the increase in numbers in countries like South Africa and Botswana appears small, it is actually a large increase by percentage.

**Fig. 3:**
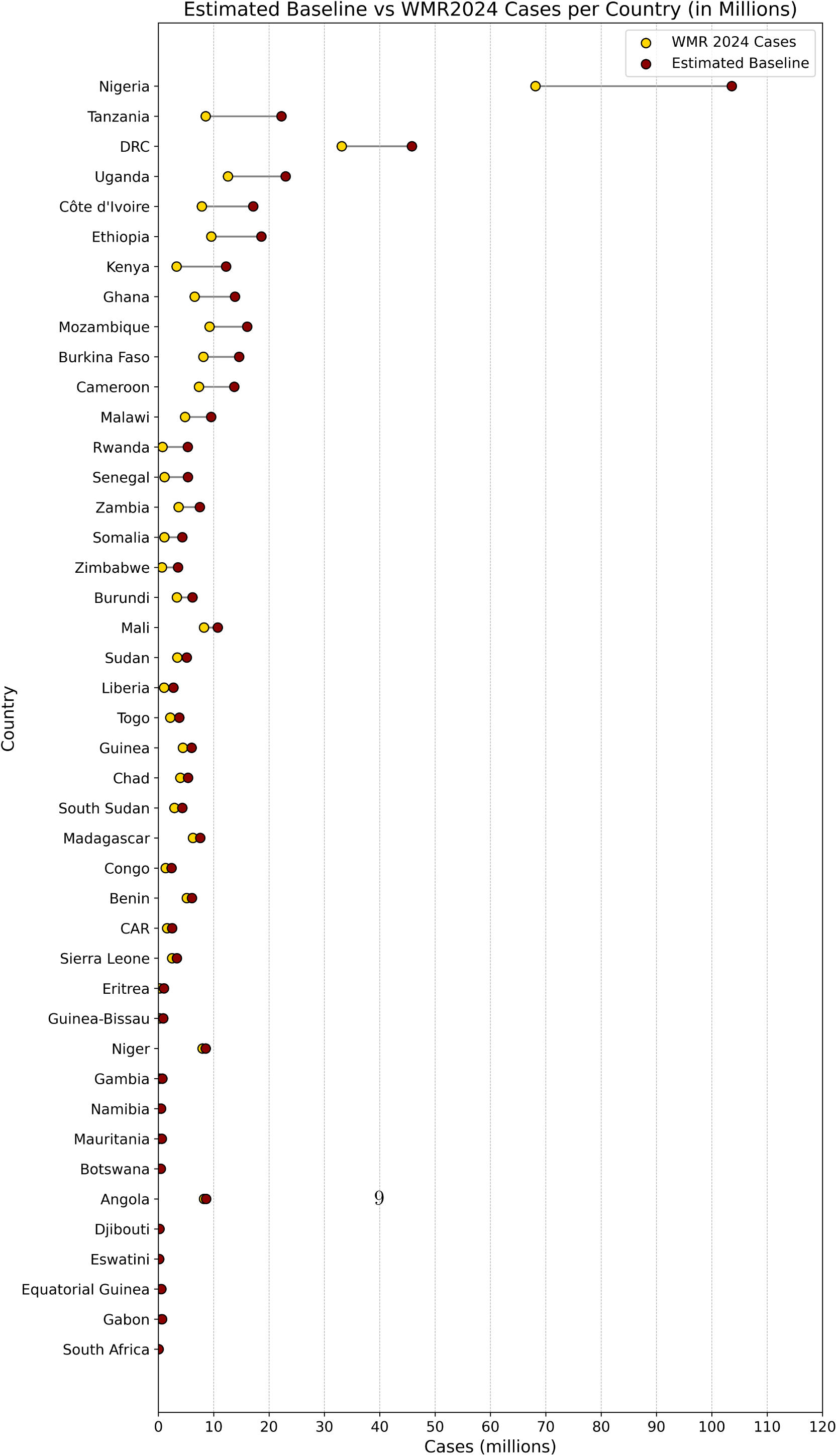
Estimated baseline numbers vs World Malaria Report 2024 numbers for the same population denominator

**Table 1:** Estimated Cases SSA Compared with WMR 2024.

| Country | Population<br>Denominator<br>2023<br>(Millions) | Estimated<br>Baseline<br>(2.5%)LI<br>(Millions) | Estimated<br>Baseline<br>Cases<br>(Millions) | Estimated<br>Baseline<br>(97.5%)UI<br>(Millions) | % Increase<br>From<br>WMR 2024 | Increase<br>(Millions)<br>from<br>WMR 2024 |
| --- | --- | --- | --- | --- | --- | --- |
| Angola | 36.71 | 6.24 | 8.63 | 11.51 | 39.52 | 3.26 |
| Benin | 14.67 | 3.36 | 6.06 | 9.11 | 77.58 | 3.98 |
| Botswana | 2.42 | 0.32 | 0.44 | 0.58 | 72400 | 0.4392 |
| Burkina Faso | 24.82 | 11.35 | 14.59 | 18.21 | 123.71 | 10.07 |
| Burundi | 14.60 | 3.46 | 6.17 | 8.84 | 163.88 | 5.49 |
| Cameroon | 34.37 | 8.38 | 13.70 | 19.23 | 161.99 | 11.89 |
| CAR | 5.78 | 1.24 | 2.49 | 3.50 | 122.93 | 1.93 |
| Chad | 19.94 | 4.00 | 5.37 | 7.03 | 77.98 | 3.08 |
| Congo | 6.03 | 1.72 | 2.38 | 3.15 | 136.84 | 1.82 |
| Côte d'Ivoire | 35.27 | 12.96 | 17.14 | 21.67 | 176.41 | 13.83 |
| DRC | 97.64 | 31.32 | 45.82 | 59.76 | 80.33 | 26.62 |
| Djibouti | 1.18 | 0.12 | 0.20 | 0.31 | 675 | 0.19 |
| Equatorial Guinea | 1.55 | 0.40 | 0.55 | 0.73 | 69.77 | 0.3 |
| Eritrea | 7.09 | 0.72 | 1.01 | 1.39 | 531.82 | 1.00 |
| Eswatini | 1.18 | 0.10 | 0.14 | 0.19 | 37900 | 0.1395 |
| Ethiopia | 116.02 | 13.77 | 18.60 | 24.44 | 155.65 | 14.88 |
| Gabon | 1.82 | 0.37 | 0.67 | 0.96 | 68.42 | 0.39 |
| Gambia | 2.42 | 0.51 | 0.71 | 0.91 | 279.17 | 0.67 |
| Ghana | 32.94 | 9.13 | 13.84 | 18.33 | 179.85 | 11.78 |
| Guinea | 14.44 | 3.30 | 6.00 | 8.77 | 97.97 | 4.34 |
| Guinea-Bissau | 2.10 | 0.46 | 0.87 | 1.35 | 486.96 | 0.42 |
| Kenya | 51.89 | 8.64 | 12.23 | 16.58 | 402.42 | 11.28 |
| Liberia | 5.50 | 2.09 | 2.72 | 3.38 | 228.15 | 2.35 |
| Madagascar | 30.66 | 4.17 | 5.56 | 7.18 | 15.07 | 0.94 |
| Malawi | 21.02 | 7.20 | 9.53 | 12.17 | 153.01 | 7.36 |
| Mali | 25.02 | 7.35 | 10.74 | 14.73 | 78.98 | 6.5 |
| Mauritania | 4.55 | 0.12 | 0.63 | 1.76 | 826.32 | 0.57 |
| Mozambique | 33.05 | 11.61 | 16.04 | 20.47 | 121.06 | 11.21 |
| Namibia | 3.02 | 0.33 | 0.49 | 0.72 | 3500 | 0.45 |
| Niger | 28.13 | 3.06 | 7.55 | 14.08 | 56.8 | 5.1 |
| Nigeria | 252.11 | 60.39 | 103.61 | 147.40 | 116.32 | 79.26 |
| Rwanda | 14.25 | 4.00 | 5.31 | 6.91 | 821.33 | 5.16 |
| Senegal | 18.11 | 3.23 | 5.34 | 8.00 | 614.29 | 5.22 |
| Sierra Leone | 7.60 | 1.70 | 3.34 | 4.79 | 93.15 | 2.31 |
| Somalia | 22.74 | 3.19 | 4.33 | 5.86 | 442.6 | 3.78 |
| South Africa | 49.32 | 0.032 | 0.047 | 0.068 | 1260 | 0.043 |
| South Sudan | 10.77 | 2.70 | 4.32 | 6.19 | 112.71 | 3.28 |

**Table 1 – continued**
| Country | Population Denominator (Millions) | Estimated Baseline LI (Millions) | Estimated Baseline Cases (Millions) | Estimated Baseline UI (Millions) | % Increase From WMR 2024 | Increase (Millions) from WMR 2024 |
| --- | --- | --- | --- | --- | --- | --- |
| Sudan | 38.49 | 3.81 | 5.14 | 6.66 | 95.31 | 3.25 |
| Tanzania | 65.83 | 13.32 | 22.24 | 32.03 | 274.62 | 13.48 |
| Togo | 8.67 | 2.19 | 3.77 | 5.36 | 150.47 | 3.22 |
| Uganda | 46.73 | 17.06 | 22.99 | 30.47 | 142.4 | 17.9 |
| Zambia | 21.55 | 4.46 | 7.49 | 10.62 | 190.16 | 6.96 |
| Zimbabwe | 15.85 | 1.40 | 3.56 | 7.15 | 1017 | 3.21 |
| SSA | 1248.42 | 275.29 | 422.36 | 582.54 | 131.32 | 330.71 |

### World Health Organisation Region Estimates

The WHO World Malaria Report categorises SSA countries into 4 broad regions: West Africa, Central Africa, Countries with High Transmission in East and Southern Africa, and Countries with Low Transmission in East and Southern Africa. Figure 5 shows the estimated incidence rates of malaria according to these WHO regions. We broadly see that if interventions stopped and there was a return to baseline, West Africa and Central Africa would be the most affected. Several countries which currently have low transmission rates will see an increase in malaria cases (Figure 5). In West Africa, the highest incidence of the proportion of the population infected would be found in Burkina Faso: 0.588 (0.457 - 0.734); Ĉote d’Ivoire: 0.486 (0.366 - 0.614), and in particular in the districts of Comoe, Lacs, Woroba, Sassandra-Marahoue and Savanes; Ghana: 0.42 (0.277 - 0.375), mainly in Ahafo, Bono, Northern, Savannah, Upeer East and West, and Oti districts; and Nigeria: 0.411 (0.30 - 0.585), with Kaduna, Oyo, Nassarawa, Akwa Ibom and Imo regions showing the highest spread. Central Africa shows extensive hotspots, particularly in DRC: 0.469 (0.321 - 0.613), highest in the Bas-Uele, Equateur, Haut-Uele, Ituri, Kwilu, Kasai Central, Mongala, Sud-Ubangi Tshopo and Tshuapa regions, Central African Republic: 0.431 (0.213 - 0.606), with Basse-Kotto, Ombella M’Poko, Lobaye regions among the highest in the country, and Cameroon: 0.399 (0.244 - 0.56), with Sud and Centre among the highest malaria regions. The countries around Lake Victoria-Uganda: 0.492 (0.365 - 0.652), Burundi: 0.422 (0.457 - 0.734), Tanzania 0.338 (0.203 - 0.469), as well as the coastal regions (Maputo, Cabo Delgado, Zambezia provinces in Mozambique: 0.486 (0.351 - 0.62)) and islands (Penba island in Tanzania, Taosmasina and Fianarantsoa in eastern Madagascar: 0.411 (0.297 - 0.527)) show very high return to baseline incidence. With minimal interventions, countries with low incidence rates such as Namibia: 0.164 (0.11 - 0.239), Botswana: 0.183 (0.134 - 0.0.241), and Eswatini: 0.118 (0.085 - 0.163) would rise to a higher baseline incidence.

According to WMR 2024, in West Africa, there were 15 endemic countries, with an estimated 124 million cases. We estimate that without interventions, a return to baseline malaria for these countries could lead to 197 million cases, with the highest number of cases in Nigeria (103 million), followed by Ĉote d’Ivoire (17 million) Burkina Faso (15 million), Ghana (14 million), and Mali (11 million), accounting for more than 80% of the cases from this region (Figure 4). This is a change from WMR 2024, with cases in baseline Ghana replacing Niger from WMR 2024. West Africa as a region would account for almost half the cases from SSA with 46.7%. In Central Africa, there were an estimated 60 million cases, according to WMR 2024, with 10 endemic countries. If malaria returns to a baseline without interventions, we estimate that this region will have 86 million cases (without accounting for Sao Tome and Principe), with DRC (46 million), Cameroon (14 million) and Angola (9 million) accounting for over 80% of the cases in this region. In countries with high transmission in East and Southern Africa, according to WMR 2024, there were 11 countries at high risk of malaria, with an estimated 62 million cases. However, in a scenario where interventions cease, we estimate that the number of cases in these countries would increase by more than 100% to 128 million. This is the region which would be most affected if interventions ceased for a number of years. Three countries, Uganda (23 million), Tanzania (22 million) and Ethiopia (19 million), would account for more than 50% of the cases in this region, with Tanzania replacing Mozambique in the set in WMR 2024. The countries in this region are susceptible to a higher amount of mosquito breeding grounds, likely attributed to the Lake Victoria basin that can support malaria transmission all year round, as well as other water bodies like Lake Malawi, Lake Tanganyika, and numerous freshwater lakes in the Great Rift Valley. The prevalence of cyclones and heavy rains, and flooding in this region also leads to breeding sites for mosquitoes [48]. And finally, in countries with low transmission in East and Southern Africa, WMR 2024 reported 267 thousand cases. We estimate 2 million cases (without Comoros) if interventions were stopped, representing a huge increase, and further demonstrating the importance of timely interventions. In this region, Eswatini (1 million) would account for 50% of the cases, with Namibia and Botswana accounting for 43% of the cases.

**Fig. 4:**
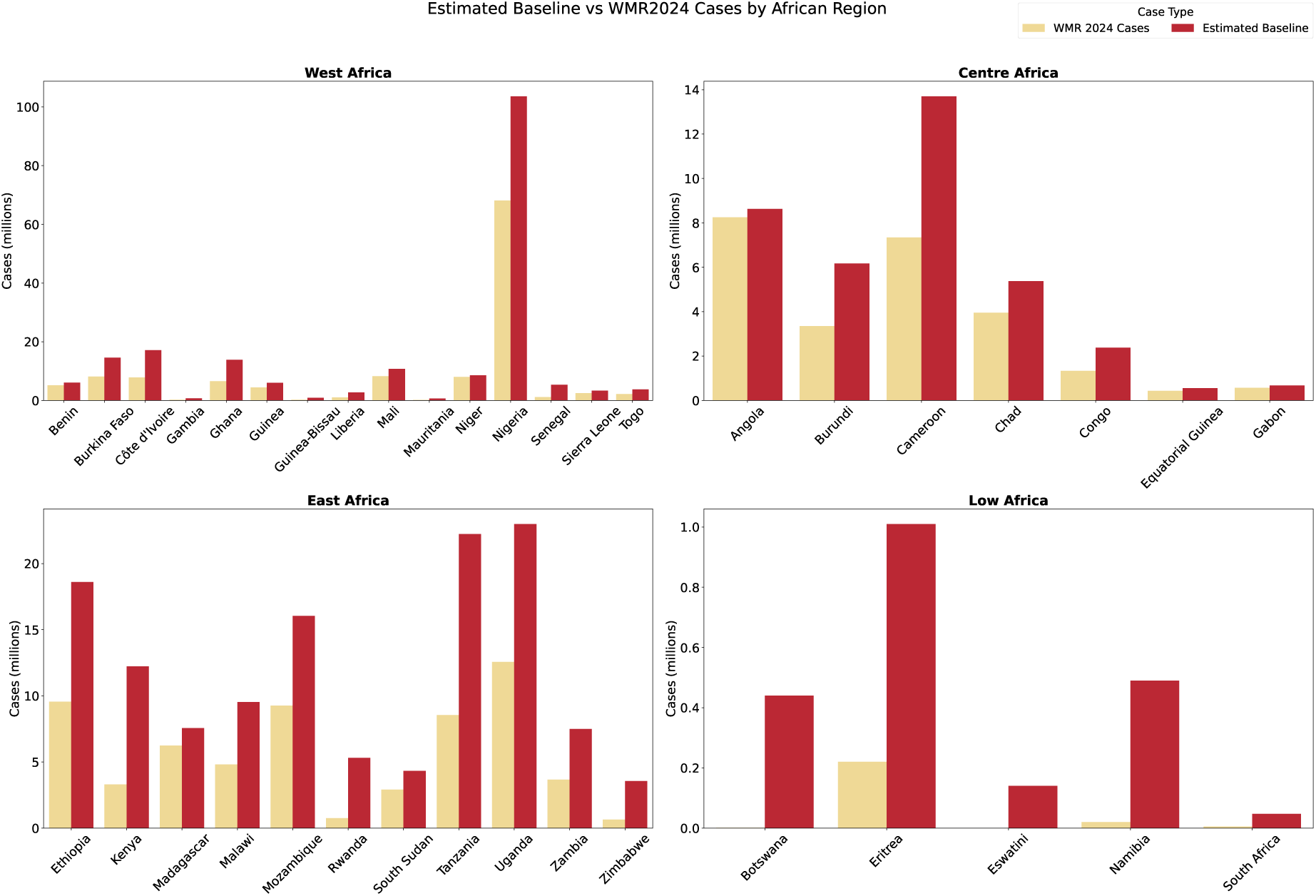
Estimated baseline numbers vs World Malaria Report 2024 numbers for WHO Regions. Top Left: West Africa. Top Right: Central Africa. Bottom Left: Countries with High Transmission in East and Southern Africa. Bottom Right: Countries with Low Transmission in East and Southern Africa.

**Fig. 5:**
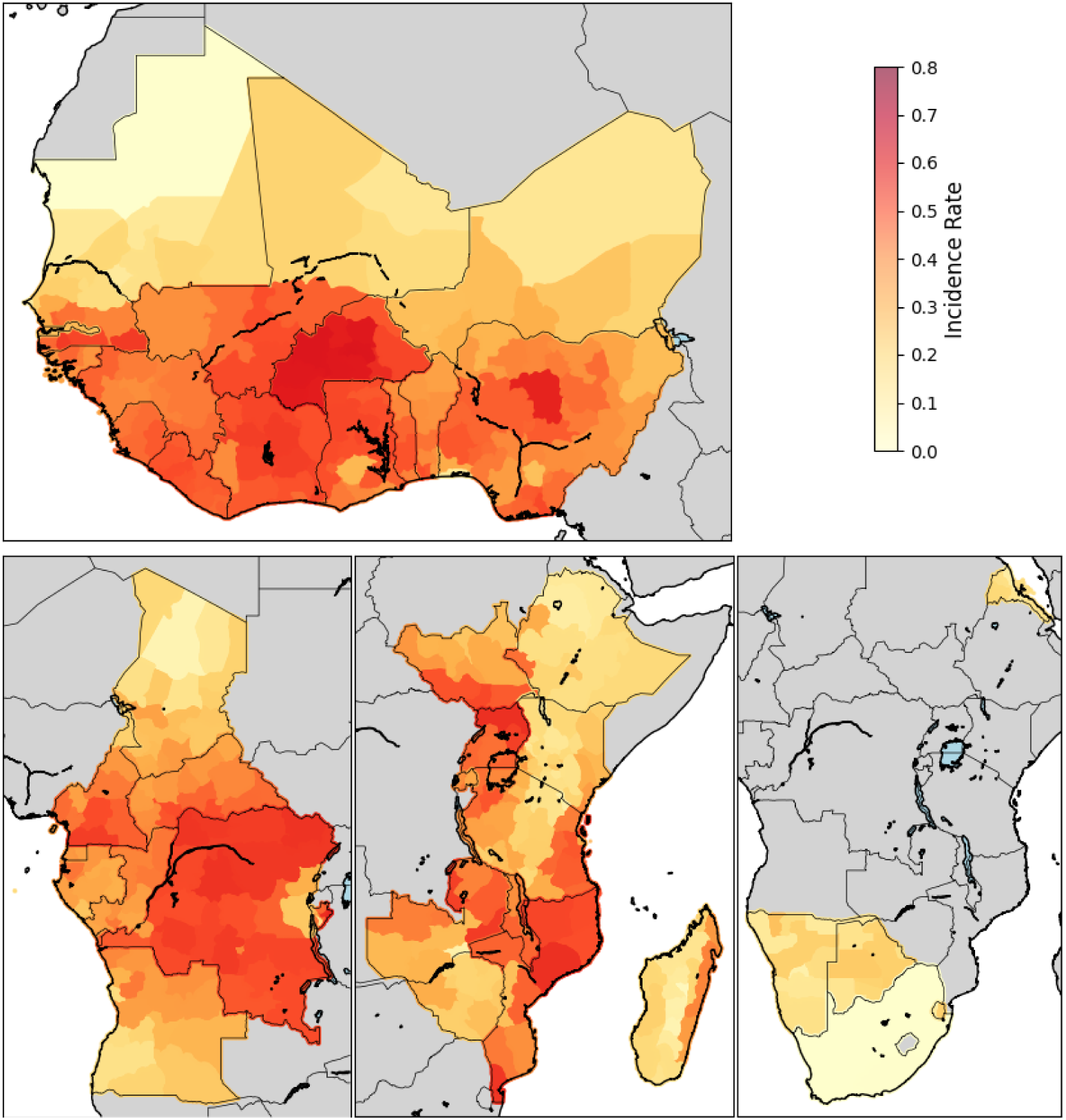
Estimated baseline malaria incidence by WHO region. Top: West Africa. Bottom Left: Central Africa. Bottom Centre: High-Transmission Countries in East and Southern Africa. Bottom Right: Low Transmission Countries in East and Southern Africa.

To summarise, in Western Africa, we estimate that malaria rates would be high throughout the region except in Mauritania, with Burkina Faso, Liberia and Ĉote d’Ivoire showing the highest transmission rates. In Central Africa, DRC, Central African Republic, and Burundi report the highest estimated malaria rate. In countries with high transmission rates in East and Southern Africa, Uganda, Mozambique, and Malawi show a high baseline transmission rate compared to the rate without interventions, while in countries with low transmission rates in the same region, all countries would show a remarkable increase in malaria transmission compared to WMR2024 numbers with interventions. The countries with high transmission in eastern and southern Africa would be the ones with the highest increase in malaria in SSA if interventions ceased.

### Estimated Baseline for High Burden to High Impact (HBHI) Countries

In 2024, the WHO classified 11 countries as high burden to high impact countries (HBHI) for malaria: Burkina Faso, Cameroon, the Democratic Republic of the Congo, Ghana, Mali, Mozambique, the Niger, Nigeria, Uganda, Sudan and the United Republic of Tanzania. These countries were responsible for 70% of global malaria cases worldwide. This makes interventions and malaria funding in these countries even more crucial. We estimate that all these countries will return to very high malaria incidence if malaria interventions were stopped, with a per cent increase in cases ranging from 56% (Niger) to as high as 274% in Ghana (Figure 6). Countries like Mali, Niger and Sudan could be replaced by Ĉote d’Ivoire and Ethiopia in the HBHI country list because of higher malaria burden. The total estimated cases from this group would increase from 173 million from WMR 2024 to 356 million, even with 1 less country in the potential HBHI group, showing a rise of. From the latest numbers from WMR 2024, Nigeria accounted for 40% of estimated malaria cases within the 10 HBHI countries, while DRC accounted for 20% of the cases in the same group. With our estimates, if all interventions are ceased, these countries would only account for 29% and 13% of the HBHI cases respectively. However, the case numbers would rise far more for the other countries in the group. Without interventions, key challenges like access to health care, ongoing conflicts emergencies and disasters, and insufficient funding would pose a significant risk towards the explosion in malaria cases.

**Fig. 6:**
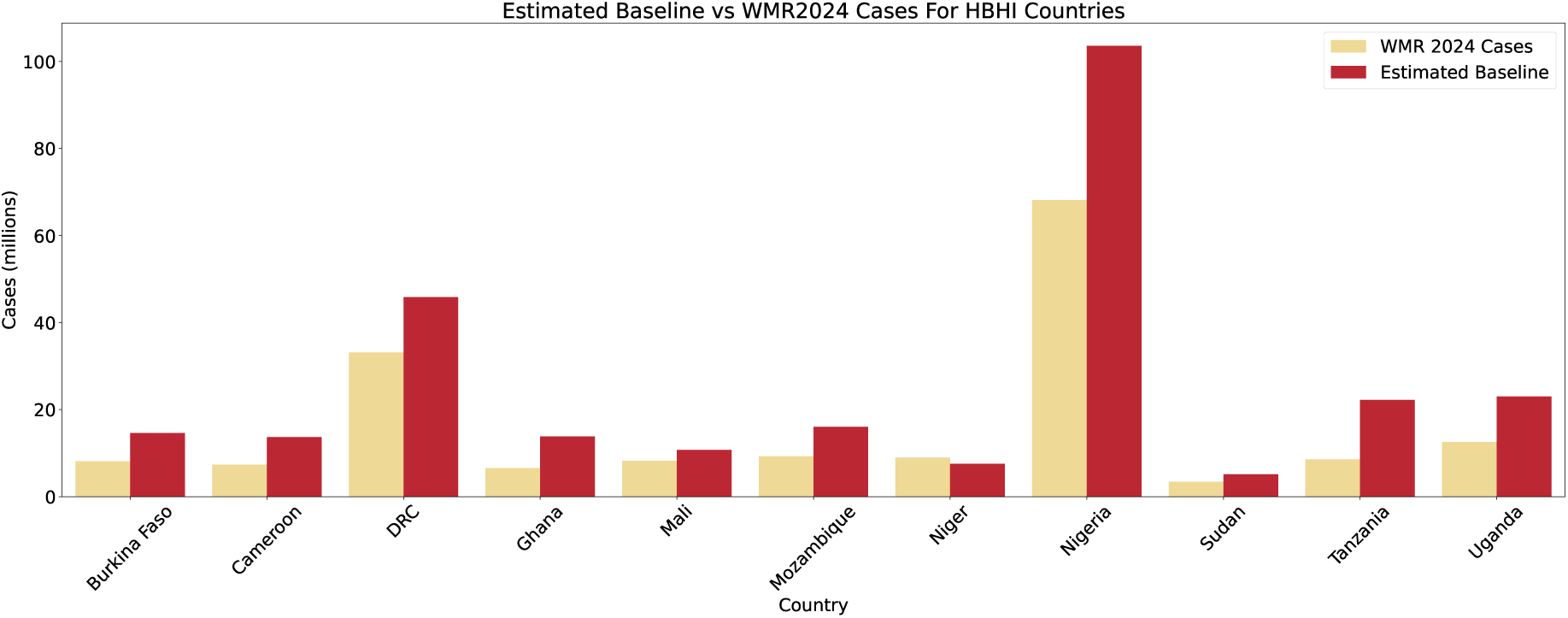
Estimated Baseline Cases compared with WMR 2024 for 11 current HBHI Countries. We see a big rise in cases in Nigeria, with around 80 million additional cases, followed by the Democratic Republic of the Congo with 265 million additional cases, while countries like Burkina Faso, Ghana, Mozambique and Cameroon show around 10-15 million increase.

### Area of coverage per results affects final predictions

We find that satellite images corresponding to a 10km x 10km area tend to offer better performance for predicting malaria compared to those covering a 5km x 5km area. There are several factors which may contribute to this performance gap. First, the 10*km*^2^ images might better account for land heterogeneity. In contrast, 5*km*^2^ images may capture only homogeneous land types, such as just forests or just deserts, without reflecting a mix of land-use types. Given the vast ecosystems in SSA, a 25km2 area may be insufficient to represent diverse land-use patterns. By averaging out the heterogeneity, the 10km images provide a more mixed signal, which could be more useful for malaria prediction. The 10*km*^2^ scale is more effective at capturing global patterns. Smaller patches, like those from 5*km*^2^ images, might cause models to focus too much on local patterns and miss broader contextual relationships. For instance, in a region like North West Nigeria, a 5*km*^2^ image might only cover desert, leading to a malaria prediction close to zero. However, a 10*km*^2^ image from the same central coordinates would also include surrounding regions, such as lakes and green areas, resulting in a malaria prediction that is more accurate, having more information about the land use types, and thus, capturing mosquito and malaria spread better. Additionally, the increased variability in the 10*km*^2^ images could be advantageous. When using contrastive learning, which relies on comparing similarities, a lot of homogeneity between patches of 5*km*^2^ (such as numerous pure desert examples) may lead to less reliable embeddings and reduced generalizability. A larger area, like 10*km*^2^, may provide a more balanced set of examples that include mixed land uses, leading to more reliable predictions. This broader context can improve model performance by offering a more comprehensive view of the environment.

## Discussion

Here, we establish a baseline malaria map, with the goal of studying the hypothetical malaria incidence rate in populations if interventions ceased but all else remained the same. Despite significant intervention efforts and considerable funding directed at prevention, diagnosis, and treatment, malaria continues to pose a serious global health challenge. The disease still accounts for millions of cases and over half a million deaths annually, predominantly impacting vulnerable populations in endemic regions. This persistence of high morbidity and mortality underlines the critical role of ongoing interventions; without them, the scenario would undoubtedly be severe, with infection rates and death tolls rising to levels higher than those recorded today. Compared to the WHO’s World Malaria Report 2024, our analyses suggest much higher estimated malaria cases, with a difference as high as 79 million cases in countries with high malaria transmission, like the Democratic Republic of Congo and Ĉote d’Ivoire. Even in countries currently experiencing low transmission of malaria, for example, South Africa, Namibia, Eswatini, and Eritrea, there are regions where incidence rates rise back to high levels, with the difference in estimated cases as high as 100-fold. Our baseline map highlights just how profound the changes in the malaria landscape have been over time and the risk inherent in the slowing of that progress.

In the pursuit of more accurate and efficient malaria estimation, the integration of satellite imagery and deep learning techniques represents the methodological state-of-the-art. By using satellite data, we can extract valuable information about environmental factors that influence malaria transmission, such as land cover, water bodies, and climate variables. Our model can analyse this data to identify patterns and make predictions about malaria risk. This method not only enhances the precision of current malaria estimates but also in malaria cases if interventions were halted. Our baseline estimates were created by embracing advanced methodologies and better leveraging available data resources. This approach offers a novel way or improving contemporary malaria burden estimates through enhanced generation and evaluation of covariates that correlate with malaria prevalence. Furthermore, the use of contrastive learning over traditional covariates gives a two-fold benefit; satellite imagery is available even in periods when it might not be possible to conduct surveys, and remote measurements can collect data from regions that are inaccessible due to various factors, including war, political instability, and pandemics. Second, Satellite images can accurately capture environmental features even over large distances, as seen by the experiment with the environmental vegetation index (Supplementary Figure 9).

The study’s predictions of malaria in areas without interventions are based on two fundamental assumptions. Firstly, it is assumed that changes in malaria prevalence over time are primarily due to interventions, such as insecticide-treated nets or improved healthcare access. This assumption implies that observed variations in malaria cases are largely attributable to these specific control measures. Secondly, the study assumes that other factors influencing malaria transmission, such as climate, ecological conditions, and socio-economic factors, remain relatively constant over the study period. This simplification allows the model to focus on the impact of interventions. However, we acknowledge that these other factors can independently affect malaria risk, and changes in these factors, as captured in satellite images from a later period, must be carefully considered to ensure accurate predictions in the absence of interventions. It is therefore a limitation of our study. As we have previously highlighted, this assumption is simplifying but is illustrative of the consequences of a rollback of interventions to pre-millennium development goals levels. In most countries, our estimates represent a plausible worst-case scenario for malaria if interventions were removed because other factors related to malaria transmission, such as housing quality and national healthcare systems, have generally improved since 2000. However, the trajectory of improvement has been highly heterogeneous across SSA (even within country-regions), with some areas backsliding in response to factors such as conflict and political instability. As such, for many locations, our baseline estimates will accurately reflect the impact of intervention removal, given a few critical drivers of malaria transmission between 2000-2025.

Based on our findings, we give some suggestions in terms of tackling malaria in the future. The impact of interventions is evident, as low-transmission countries would have experienced an estimated increase of 1.73 million cases without any interventions, compared to the latest numbers. Low-transmission countries have reached a state of low malaria transmission because of years of intervention and funding efforts. Without these efforts, any inroads made towards malaria elimination would start to erode. There are a lot of challenges involved in accessing remote areas for collecting survey data, and these surveys are often expensive and rely entirely on external funding. The use of satellite imagery and deep learning can be a low-cost alternative or addition to these surveys. Additionally, there are often missing covariates for some regions because of the reasons above, and satellite images can supplement these missing covariates. Finally, much has been said about the rising temperatures due to climate change and its impact on malaria transmission levels [5, 49]. Our analyses here indicate that the risks associated with halting interventions are more immediate and significant than those posed by long term climate change. While climate change may influence malaria transmission, the effects of increased temperatures in SSA are complex and may not uniformly increase transmission. Therefore, we recommend that continuing and improving intervention strategies, particularly in the HBHI countries, should be the top priority.

### 1 Data Sources, Materials and Methods

Three different types of data sources were used: satellite images, *P. falciparum* infection prevalence data collected during household surveys, and a set of covariates developed by [50] that are used to build a dataset with which to compare our contrastive deep learning method.

#### Remote-Sensed Satellite Imagery

281,846 Landsat 8 satellite images were obtained via Google Earth Engine [43, 51] for sub-Saharan countries over the period of 2013-2022. Each image covers a 10×10 km footprint, consists of 333×333 pixels with a spatial resolution of 30 meters, and each pixel contains eight spectral bands. For the purposes of this project, we used the Red-Green-Blue (RGB) bands. A single composite image was created to represent images from one year from a particular 10×10 km area.

The images are preprocessed using radiometric and geometric calibration to correct for sensor and platform errors. Radiometric calibration involves converting the raw pixel values to physical units, while geometric calibration involves correcting for any distortions caused by the satellite’s orbit. The preprocessed images are then ready for use in various applications such as land cover mapping, change detection, and vegetation monitoring [43].

The imagery dataset is geographically diverse, covering various natural environments (e.g., forests, deserts, rivers) and human-made infrastructure (e.g. urban centres, road networks) (Figure 7).

**Fig. 7:**
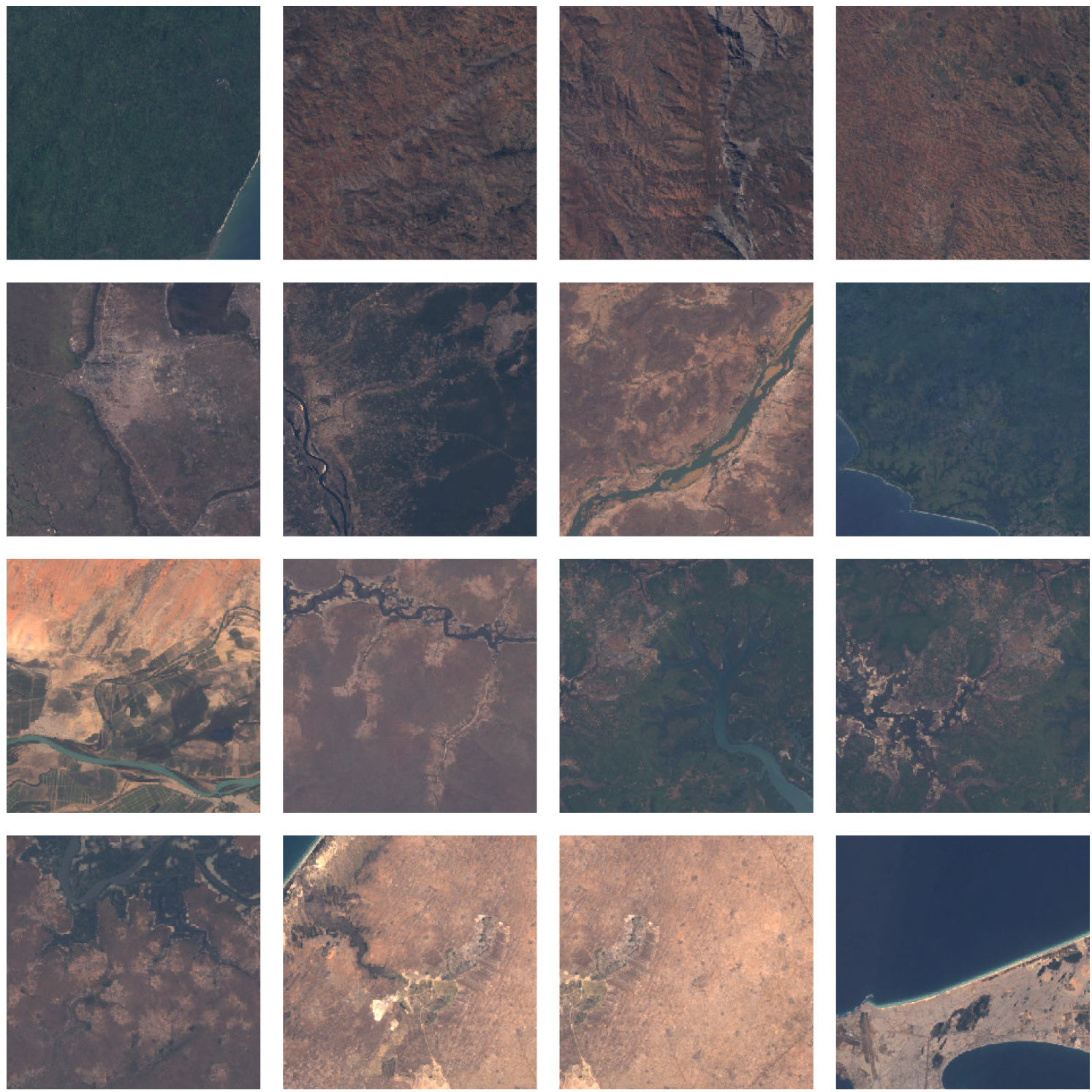
Images acquired by Landsat 8

The use of satellite imagery acts as a proxy to covariate data- for example, land cover, vegetation indices, elevation, and built-up environments like road densities, urban networks, and population centres. All of this information can be extracted from the satellite images using deep learning models to supplement covariate sources [52–56].

The ideal way to approach this analysis would be to train the classification model using remotely sensed imagery collected contemporaneously with the survey points and then make predictions by applying the model parameters to recently collected and identically structured imagery. However, despite the depth of the Landsat 30-meter archive, which extends back to the 1980s, the availability, quality, and consistency of data from the Landsat 4, 5, and 7 sensors are lower than that of the Landsat 8 data currently being collected. However, previous satellites also have issues with data collection and processing, like scanline issues in Landsat 7 scenes [57–60]. Given the tremendous volume of images being processed for this research, introducing additional preprocessing steps to integrate earlier Landsat data was not feasible. Instead, because the desired outcome of this work is a map of the present day, hypothetical *PfPr* in the absence of interventions, we elected to train and apply the model to only modern imagery, thereby introducing discontinuities for survey points collected in areas that have experienced landcover change since the survey was conducted. While such discontinuities introduce noise into the linkage between response and predictor variables, the large number of survey points and typically gradual nature of landcover change ensure sufficient signal for the model to perform accurate predictions.

#### Survey Data

Part of the malaria survey data came from the Service Provision Assessment (SPA) data from the Demographic Health Surveys (DHS) [61] conducted by the United States Agency for International Development (USAID) between 2000 and 2008 for malaria infections related to *Plasmodium falciparum*. We also used data from various World Malaria Reports. The outcome variable consisted of observations of *P. falciparum* infection prevalence (*PfPr*), describing the percentage of individuals (usually children under 5) who tested positive relative to all children within a survey cluster. The position of each cluster is captured with geographic coordinates, with exact latitude and longitude displaced to preserve anonymity [62]. The full survey dataset was based on Bhatt 2015 and extracted from Pfeffer 2018 [3, 63] and consisted of 53,750 points from 45 SSA countries. These data, which are a mix of biased surveys[34] and cross-sectional household surveys contain several limitations. First, they are spaced unevenly through space and time. Second, positive malaria diagnoses from the surveys may not correspond directly to the timing of infection, as initial malaria infections could have transpired months or even years before the survey date, and malaria infections can also remain detectable for several weeks after the infection has cleared. Third, the spatial resolution of national surveys is not precise due to privacy considerations and more broadly corresponds to some point within a 10km range. This displacement is not prohibitive for the analysis because, despite using 30-meter predictor data, ultimately, the combined properties of pixels comprising the surrounding landscape are used to estimate malaria burden. Finally, they correspond to a diverse range of age types of the population with different malaria diagnostic types. These limitations are mitigated by post adjustment of the prevalence data[30, 31].

#### Data on baseline malaria prevalence

Implicit in our approach are the following assumptions: First, that changes in parasite rate over time have been primarily driven by the large-scale implementation of interventions across SSA. This is supported by a number of studies [3, 64–84], and no other compelling explanation has been provided to explain the major continental shifts in parasite rate over time. The second major assumption is that the ecological niche of malaria is relatively stable over time, especially over a time scale of a few decades. This is undoubtedly a simplifying assumption, but we believe it is justified as a first approximation. Only 3% of the Earth contains a human settlement, and the number of new settlements emerging tends to be smaller than the enlargement of existing ones [85–87]. Moreover, the rapid population growth and urban expansion in SSA, while significant, do not necessarily imply drastic changes in the environmental features relevant to malaria prevalence. For example, the doubling of cropped area between 1975 and 2013 in West Africa was primarily driven by agricultural transformation rather than changes in the underlying environmental conditions [88]. Indeed, when examining satellite imagery for the years 2000 and 2017 for a subset of our baseline locations, we find a correlation across pixels of 0.93 (2000) to 0.91 (2017), indicating a degree of stability in the ecological environment over time. A third limitation is that our modelling framework does not account for seasonality, a factor also largely ignored in previous large-scale malaria mapping efforts[3]. This was influenced by the nature of the satellite data used, which required combining many years of data to form a reliable composite image. This choice may lead to biases as the parasite rate surveys may be sampled at different seasonal points, and the composite images may be biased to cloud-free data.

Our full prevalence dataset captures malaria observations in locations that have undergone substantial fluctuations over time due to a myriad of significant changes across SSA and, most importantly, the implementation of various interventions[3]. While it is possible to characterise these changes through mapping[3, 15], estimating the prevalence if interventions were removed requires additional modelling on a subset of the response data that had minimal intervention impacts. Previously, Bhatt et al. [3] modelled estimated malaria change using interventions and then estimated counterfactuals through simple g-computation. This approach, however, relies on an accurate characterisation of changes in intervention coverages across both space and time, which is challenging[89]. Our conceptually similar approach is to model prevalence using a subset of data points recorded before interventions were introduced (Supplementary Figure 11). This approach presents challenges, such as the necessity to establish thresholds for intervention coverages that constitute “pre-intervention”. Attempting to mitigate these complexities, we used a maximum threshold of 10% average usage of insecticide treated bed-nets over 4 years. Theoretical frameworks suggest that this threshold’s impact on prevalence remains minimal[36]. Empirical observations further support this assertion, as marginal differences are noted in the mapped outcomes for thresholds set at 5% and 15%, particularly at a regional level (Supplementary Figure 10). When considering the impact of effective treatment with an antimalarial drug, a critical consideration is that drugs primarily prevent malaria mortality rather than directly influencing the prevalence by the vector or sustained transmission control[90]. Based on existing literature, we select a threshold of 50% for effective drug treatment because in high transmission settings, increasing treatment coverage is associated with declines in mortality rate, but the impact of increasing treatment coverage on incidence is less pronounced [91]. This exclusion also helps to mitigate the impact of antimalarials. Artemisinin based combination therapies (ACTs) were substantially rolled out after 2008, and extensive drug resistance had been observed for other antimalarials (chloroquine, sulfadoxine), which were first line therapies before ACTs [92–94].

While it is challenging to obtain a fully controlled dataset that accounts for all factors such as seasonality, changing climate, urbanisation, culture, and economics, our constructed dataset offers a robust modelled estimate of the potential malaria prevalence landscape in the hypothetical absence of interventions, assuming sustained socioeconomic progress. This approach provides valuable insights despite the inherent methodological assumptions and limitations in controlling every confounding factor.

### Modelling

#### Overview

Here, we describe the main methodological steps to implement, train, and evaluate our model (Figure 1). First, we pre-train a deep convolutional neural network (CNN) on a diverse set of satellite images from SSA using a self-supervised contrastive learning technique to extract the image features. Pre-training using self-supervised learning allows deep neural networks to learn a meaningful representation from large, unlabelled datasets. In particular, contrastive learning is an efficient method to achieve this goal, as the model learns to minimise the similarity of dissimilar sample image pairs (i.e., negative pairs). Importantly, the learned features can be used for various downstream tasks using transfer learning [95, 96]. In this framework, after pre-training, embeddings for each satellite image were extracted from the pre-trained model before being used for malaria prevalence prediction for the images’ respective geographic regions.

#### Image Preprocessing

In the first step, each satellite image *V* is converted into two views, *v*1 and *v*2. These views are generated using colour and geometric augmentations. The colour augmentations included adjusting saturation, hue, contrast and brightness, while the geometric augmentations consisted of cropping, horizontal and vertical flips and rotations by 90 and 180 degrees. Compared to previous contrastive learning methods, instead of using purely random augmentations, we decided to make contrastive augmentations for each view (explained below). This was to ensure an increase in difficulty during training, as the model needs to learn similarity between 2 images which are further apart than only random augmentations, which in turn would make the model more robust to smaller perturbations in the data. Thus, both views *v*1 and *v*2 undergo both contrasting geometric and colour augmentations. The brightness of all nighttime satellite images was first increased before any other augmentations were done. The full list of augmentations can be seen in table 2.

**Table 2:** Contrasting augmentations performed on both views of the image. Some augmentations, like adjusting brightness, random crops, were applied to both views, while all other augmentations were applied contrastively for the different views. Note that all augmentations except for adjusting brightness, flips and crops were applied with a probability of 0.75.

| View 1 | View 2 | Augmentation Type | Probability distribution | Scale |
| --- | --- | --- | --- | --- |
| Both views |  |  |  |  |
| ✓ | ✓ | Random Crop | Uniform | (224 px, 224 px) |
| ✓ | ✓ | Adjust brightness | Uniform | $\pm 0.75$ ($\Delta 0.75$) |
| View 1 only |  |  |  |  |
| ✓ | × | Random rotation | Uniform | $\pm 90^\circ$ |
| ✓ | × | Random vertical flip | Uniform |  |
| ✓ | × | Adjust contrast | Uniform | $\pm 1.9$ |
| ✓ | × | Adjust saturation | Uniform | $\pm 1.1$ |
| ✓ | × | Adjust hue | Uniform | $\pm 0.01$ |
| View 2 only |  |  |  |  |
| × | ✓ | Random rotation | Uniform | $\pm 180^\circ$ |
| × | ✓ | Random horizontal flip | Uniform |  |
| × | ✓ | Adjust contrast | Uniform | $\pm 2.5$ |
| × | ✓ | Adjust saturation | Uniform | $\pm 0.75$ |
| × | ✓ | Adjust hue | Uniform | $\pm -0.01$ |

#### Pretraining

Contrastive deep learning is a technique used for unsupervised representation learning, where the goal is to learn a representation of data such that similar instances are close together in the representation space while dissimilar instances are far apart. This method enhances the performance of various tasks by contrasting samples against each other to learn common attributes between data classes and attributes that distinguish one class from another. In contrastive learning, augmenting images is a crucial step that involves creating modified versions of the original images to form positive pairs. These augmentations can include transformations such as cropping, rotating, colour jittering, and scaling. The goal is to teach the model to recognise that these augmented versions are similar to the original despite the variations introduced. This process helps the model learn invariant features that are robust to different types of distortions, thereby improving its ability to generalise to new, unseen data [97, 98].

We built our model for pretraining based on SimSiam architecture [39], a simple framework for self-supervised contrastive learning. Like many state-of-the-art contrastive methods [40–42], SimSiam-based models learn image representations by maximising the similarity between two augmented versions of a single image. The augmented *views* are then passed through an *encoder* module (usually consisting of a convolutional neural network), which outputs a *representation* of the augmented views. Subsequently, a multilayer perceptron (MLP) with three layers (the *predictor* module) is used to predict the second representation from the first representation. As a contrastive learning signal, a negative cosine similarity loss is used:

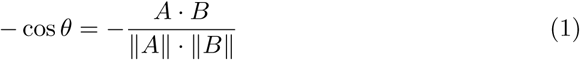

where A and B are two representation vectors, and ∥.∥ is the *ℓ*_2_ norm. Cosine similarity ranges between [−1, 1], where −1 means the vectors are opposite, 0 means they are orthogonal, and 1 means they are the same. Note that this task is equivalent to maximising the similarity between two (representations of) augmented images.

#### Encoder and predictor architecture

The two enhanced views, view v1 and view v2 are processed through a VGG-based convolutional neural network (CNN). Although ResNet-50 was used as the default encoder in the original version of SimSiam [39], we use here a smaller CNN based on the VGG architecture [99] due to the lower number of training samples (to avoid over-fitting). The input to the CNN is a 224 × 224 × 3 image, which is then fed through five convolutional blocks with Leaky ReLU activation and batch normalisation (Figure 8). Max pooling is applied after each block except for the last one, where global average pooling is applied. Finally, the output of the global average pooling operation is fed through 3 leaky ReLU-activated, and *ℓ*_2_-regularised fully-connected layers, which output the encoder representation. The choice of Leaky ReLU was empirically motivated to avoid the zero-gradient issues caused by traditional ReLU and to ensure that all neurons in the network contributed to the output [100]. The loss function, as defined in equation 1, is applied to both view v1 and view v2 as they pass through the encoder. During training, the weights are updated for view v1, while a stop-gradient technique is applied to view v2, meaning that its weights remain unchanged. Essentially, the stop-gradient method implicitly generates negative pairs.

**Fig. 8:**
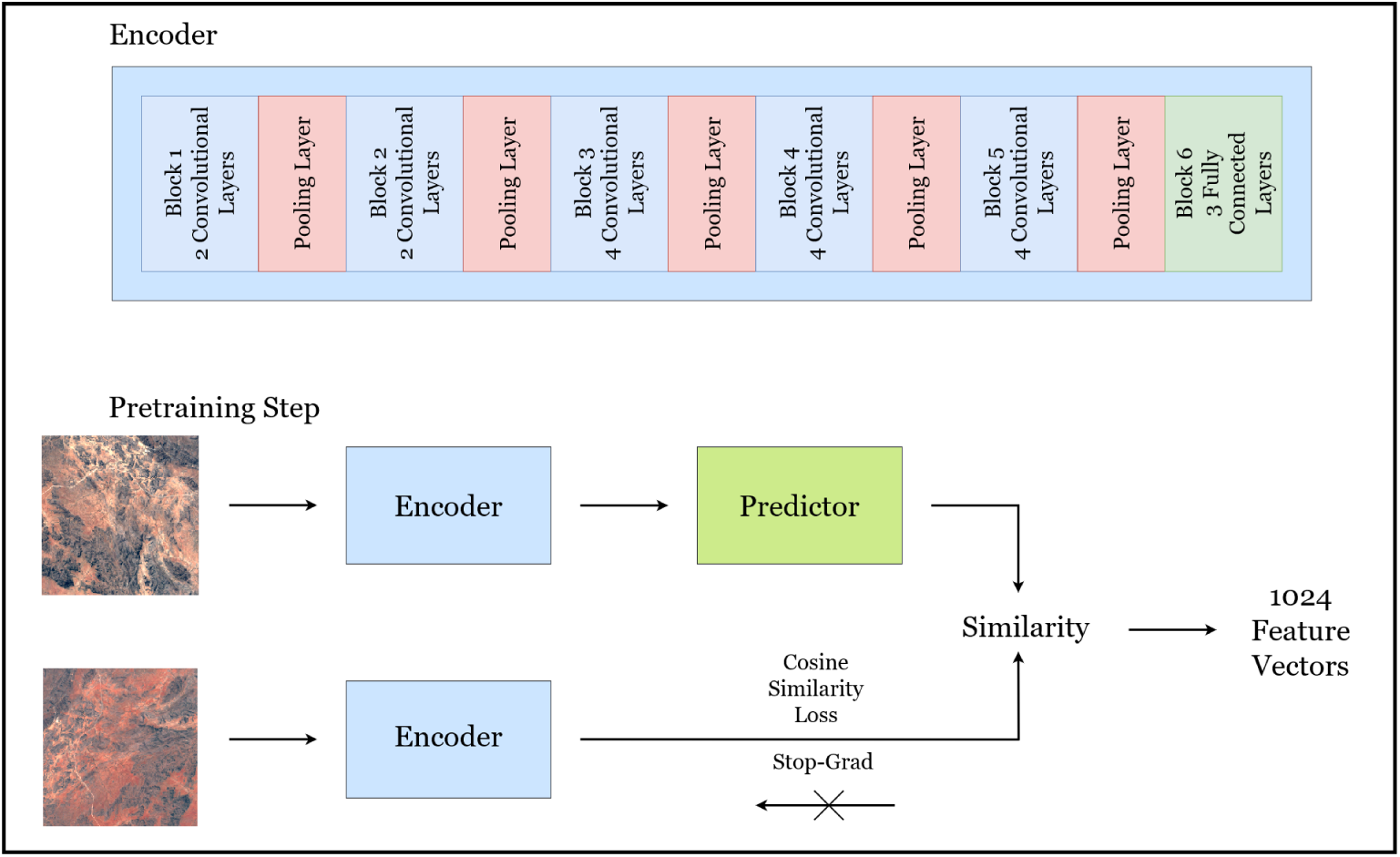
Top: Encoder architecture similar to VGG-19. Bottom: Pretraining specifics. Two augmented views, v1 and v2, from satellite image V are passed to the encoder. The weights on v1 are updated and passed to the prediction, while the weights on v2 remain static. The predictor adapts the output of v1 to align with v2 to produce a feature vector map for V.

A 3-layer multilayer perceptron (MLP) is integrated with the encoder model. The MLP’s role is to transform the output of view v1 to align with that of view v2. Since view v2’s weights are not updated, the MLP does not receive gradients from view v2’s output but does from view v1’s. Consequently, the predictor modifies view v1’s output to match view v2. The predictor architecture is a simple architecture, comprising two fully connected layers with a batch-normalised hidden layer in between. The final output of this predictor structure is a vector embedding that represents each image VV.

This first step of pretaining the contrastive deep learning model on the satellite produces 1024 feature vectors describing the land use information for all images. We experimented with changing the size of this feature set (512 and 2048) and found that 1024 gave the best experimental results.

#### Bayesian Mapping Model

We use the prevalence rate as an empirical logit, given by equation 2.

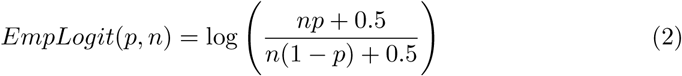

Given a basis *X* ∈ R*^n^*^×^*^m^* where, as in section X *m* is chosen as 1024, we have corresponding malaria prevalence observations 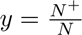 as the number of positive tests out of a sample. As noted previously these raw observations are corrected for age[30] and diagnostic type[31] yielding new *PfPr* _2-10_ observations 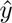. We perform the empirical logit on these observations to facilitate modelling with a Gaussian likelihood[3, 101], resulting in an estimate of malaria prevalence *y*^∗^ ∈ R. We model this prevalence using the following Bayesian hierarchical model in the Bayesian software Numpyro[45].

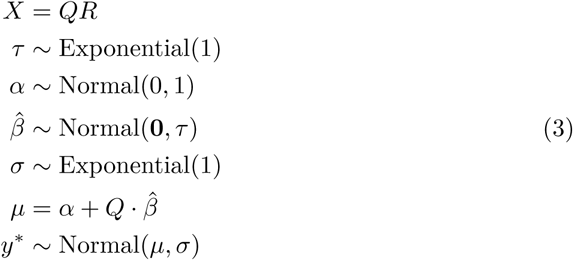

Note, as we use QR decomposition to facilitate efficient sampling, operations on the original matrix *X* can be found equivalently post-sampling by transforming the column vector 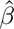

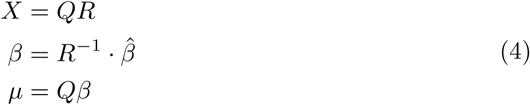

Model 3 performs Bayesian ridge regression and, therefore, attempts to mitigate overfitting from using a large number of features.

For any new prediction location, our contrastive learning model learns features *x* for any latitude and longitude within our pretraining domain. Given the sampled posterior, and collapsing all parameters into a vector *θ* = {*α, β, σ, τ* }, the predictive distribution is given by

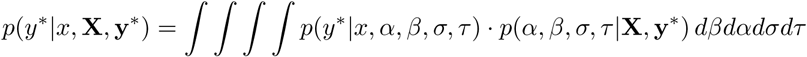

Which is approximated via Monte Carlo sampling. Note prevalence on the {0, 1} scale can be recovered through the inverse logit *ϕ*^−1^(*p*(*y*^∗^|*x,* **X**, **y**^∗^))

We generate 200 realisations by drawing different samples of coefficients from the betas. We then convert each realisation into a raster.

#### Converting Prevalence Rates to Incidence Rates

Using the ensemble model by Cameron et al., we convert the prevalence realisations into incidence rates using population rasters until 2024 [46]. We process the estimated malaria *PfPr* data by resampling it to match the resolution of population data before computing population-weighted prevalence estimates for administrative units. The population raster is cropped to match the extent of the administrative boundaries, and its zonal sum is calculated for each unit. Then, for each realisation of *PfPr*, the raster is cropped and resampled to match the population raster’s resolution to avoid distorting population counts. The prevalence rate is converted into absolute counts using the population raster, and the zonal sum of these counts is computed. The weighted prevalence is then obtained by dividing by the total population in each administrative unit. This process was repeated for all years. Finally, the zonal statistics were computed at the admin 1 level.

#### Implementation

The contrastive deep learning model was trained for 100 epochs, and each satellite image was converted into 1024 feature vectors. This implementation was performed using TensorFlow v2.4.1 [102] in Python v3.11.04. Training was performed using two GeForce RTX A6000 GPUs (Nvidia Corporation), each with an i9-9960X CPU @ 3.10GHz CPU (Intel Corporation) with 128 GB of memory and 16 cores. The downstream tasks were done using a combination of numpyro, R and Python.

Our Bayesian model shows no pathologies in posterior sampling, with no divergent transmissions and Rhat statistics very close to 1. Visual inspection of the trace also suggests a well-calibrated posterior (Supplementary Figures 12, 13 and 14).

## Data Availability

All data produced in the present study are available upon request at the different data sources, available online at https://www.dhsprogram.com/Data/, https://www.who.int/teams/global-malaria-programme/reports/world-malaria-report-2024, and https://developers.google.com/earth-engine/datasets/catalog/landsat

https://github.com/kaustubhchakradeo/Estimating-the-worst-case-scenario-for-malaria-parasite-rate-in-sub-Saharan-Africa

## Acknowledgements

KC acknowledges the grant support from William Demant Fonden (24-3395). SB acknowledges support from the Novo Nordisk Foundation via the Novo Nordisk Young Investigator Award (NNF20OC0059309), which also funds KC and AK. SB acknowledges the Danish National Research Foundation (DNRF160) through the chair grant which also supports NS. SB acknowledges support from The Eric and Wendy Schmidt Fund For Strategic Innovation via the Schmidt Polymath Award (G-22-63345). SB acknowledge funding from the MRC Centre for Global Infectious Disease Analysis (reference MR/X020258/1), funded by the UK Medical Research Council (MRC). This UK funded award is carried out in the frame of the Global Health EDCTP3 Joint Undertaking. SB is funded by the National Institute for Health and Care Research (NIHR) Health Protection Research Unit in Modelling and Health Economics, a partnership between the UK Health Security Agency, Imperial College London and LSHTM (grant code NIHR200908). DAD is supported by a Novo Nordisk Fonden Data Science Emerging Investigator grant (NNF23OC0084647). Disclaimer: ‘The views expressed are those of the author(s) and not necessarily those of the NIHR, UK Health Security Agency or the Department of Health and Social Care.’. DAD acknowledges support from the Novo Nordisk Foundation via the Emerging Data Science Investigator award (NNF23OC0084647). SM would like to acknowledge National Research Foundation via The NRF Fellowship Class of 2023 (NRF-NRFF15-2023-0010) award.

## Author Contributions

KC, SB and SM conceived and designed the study. KC linked the satellite images from Google Earth Engine to the DHS survey data. KC conducted the deep learning experiments. SB and DW completed the prevalence-to-incidence conversion. KC, AK, NS, IH, SM, PG, KB, AN, DS, SF, DD, DW and SB all reviewed the manuscript and contributed to its scientific interpretation.

## Supplementary

We present the experiments with enhanced vegetation index. When using satellite features, we find that the overall correlation and prediction power remains stable over 100 square kilometres, despite the changes in the land over time (Figure 9). We find a mean absolute error of 0.04 and a correlation of 0.93 for values in 2000 and a mean absolute error of 0.03 and a correlation of 0.91 for values in 2017.

**Fig. 9:**
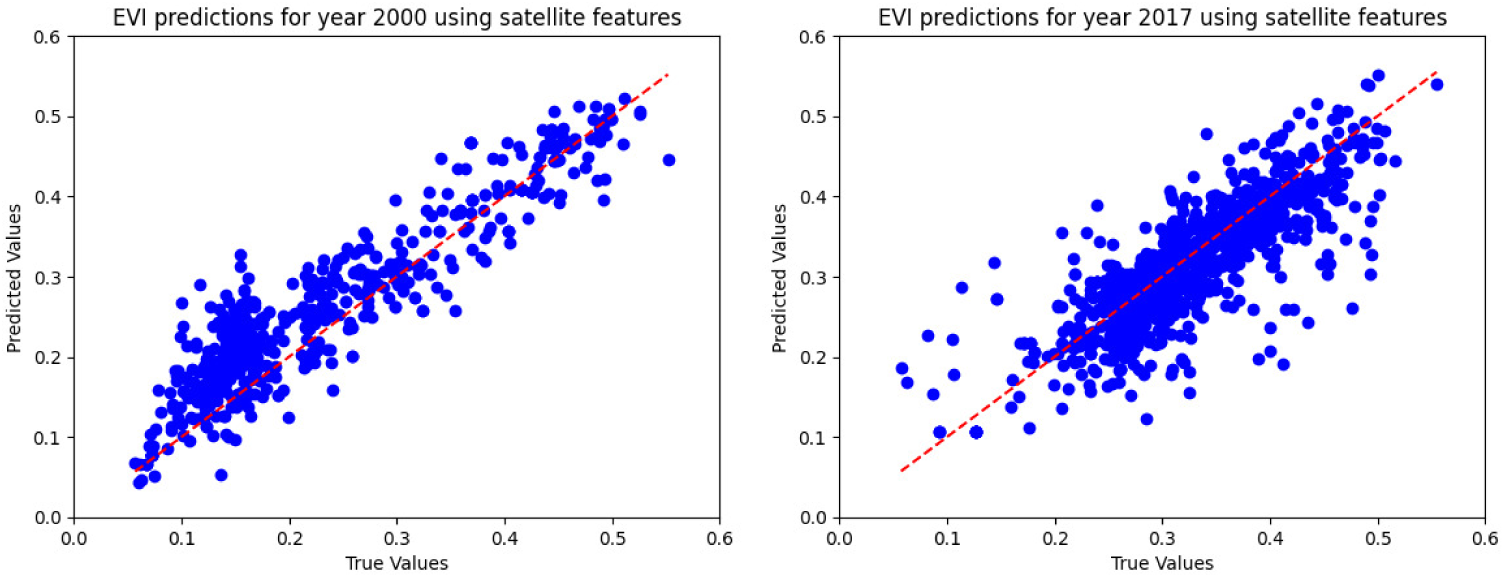
Enhanced Vegetation Index Across Years Predicted Using Satellite Features

As mentioned by Winskill et al., the effect of treatment coverage on case incidence and mortality rates. In a low transmission setting (baseline *PfPr* _2-10_: 10%), increasing treatment coverage results in reduced mortality rates and case incidence. In medium (baseline *PfPr* _2-10_: 30%) and high (baseline *PfPr* _2-10_: 60%) transmission settings, increasing treatment coverage continues to lower mortality rates. However, as transmission levels rise, the impact of increased treatment coverage on case incidence diminishes. In all scenarios, LLIN access is maintained at 25%.

**Fig. 10:**
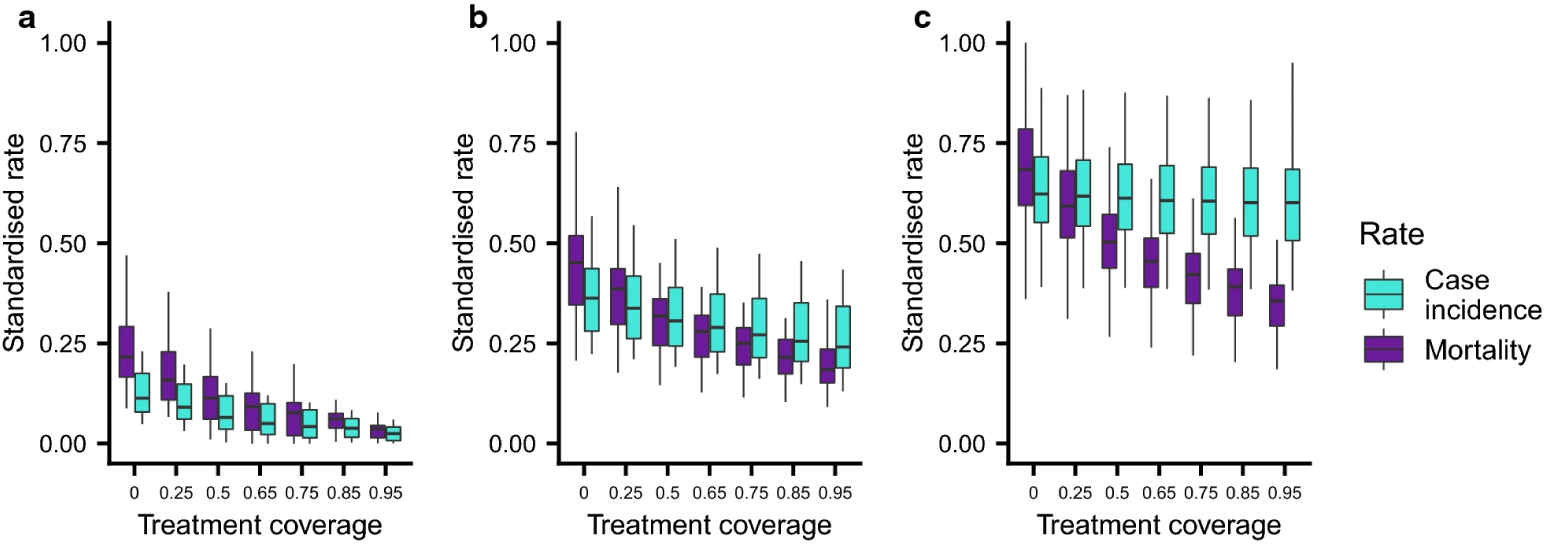
Impact of treatment coverage on incidence and mortality rates [91]

We select the baseline points with the criteria mentioned in the main body-where we select points below 50% effective drug treatment and below 10% insecticide treated bed-nets. A lot of points are from before the establishment Global Malaria Action Plan in 2008, but as interventions across countries are heterogenous, our baseline threshold map still contains recent points as well (Fig. 11).

**Fig. 11:**
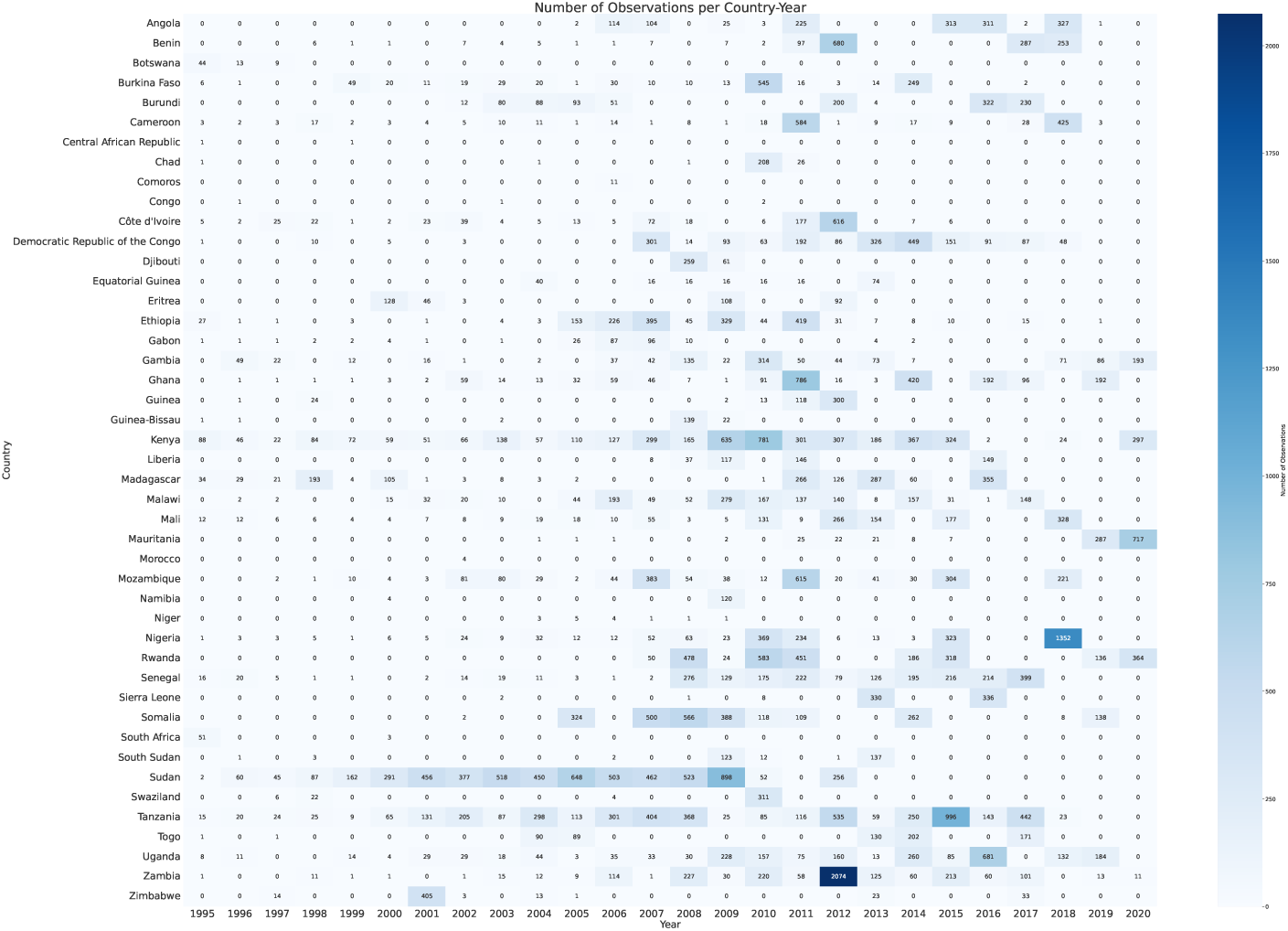
Baseline Points per Country per Year. These points satisfy the established baseline threshold criteria of less than 10% insecticide treated bed-nets and less than 50% ACTs.

We have the trace plots of the Bayesian model for selected coefficients (Figure 12). All parameters exhibit good mixing and stationarity, indicating stable sampling behaviour. No divergent transitions occurred during sampling.

**Fig. 12:**
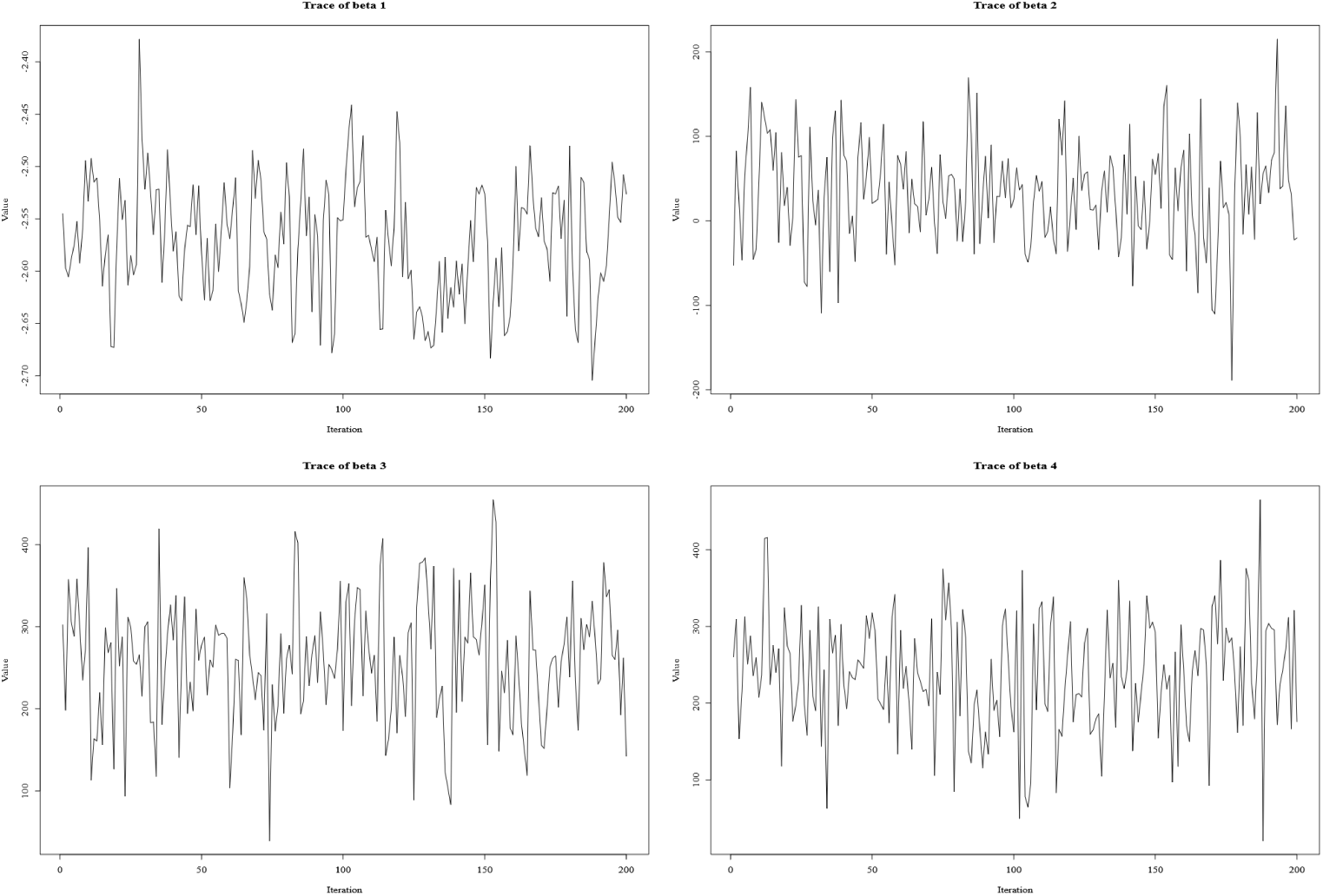
Trace Plots for Bayesian Model

We also plot posterior density plots for selected coefficients (Figure 13). All posteriors exhibit well-defined, unimodal distributions, with varying degrees of uncertainty across parameters. This indicates that the model was able to infer stable and interpretable parameter estimates.

**Fig. 13:**
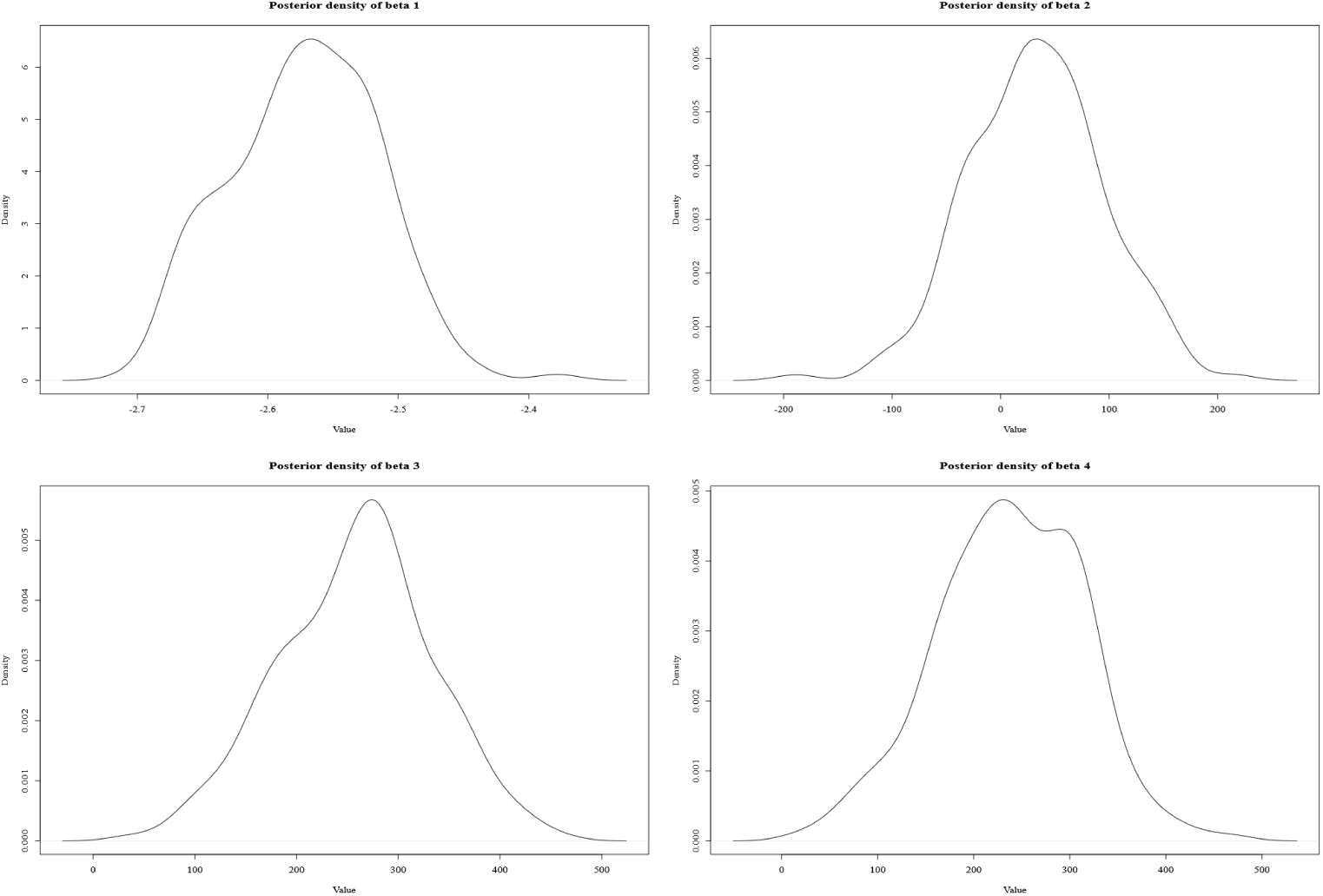
Posterior Density Plots for Bayesian Model

We show the independent effective samples that the MCMC chain is equivalent to (Figure 14). Most parameters exhibit high ESS, indicating low autocorrelation and reliable estimation. These results, along with trace and density plots, confirm good mixing and posterior stability.

**Fig. 14:**
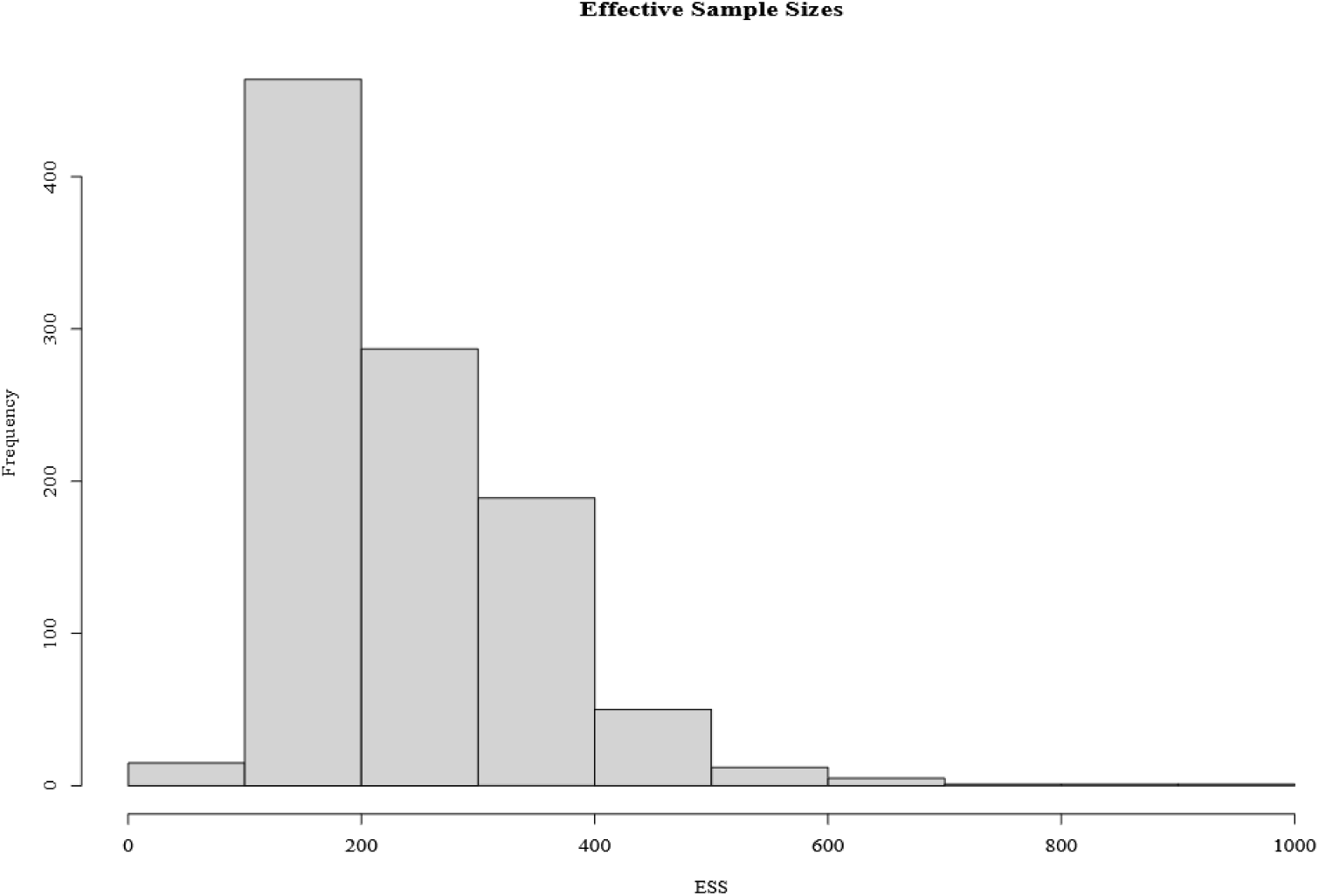
Effective Sample Sizes for Bayesian Model

We plot the estimated malaria prevalence vs empirical malaria prevalence based on the Bayesian model (Fig.15).

**Fig. 15:**
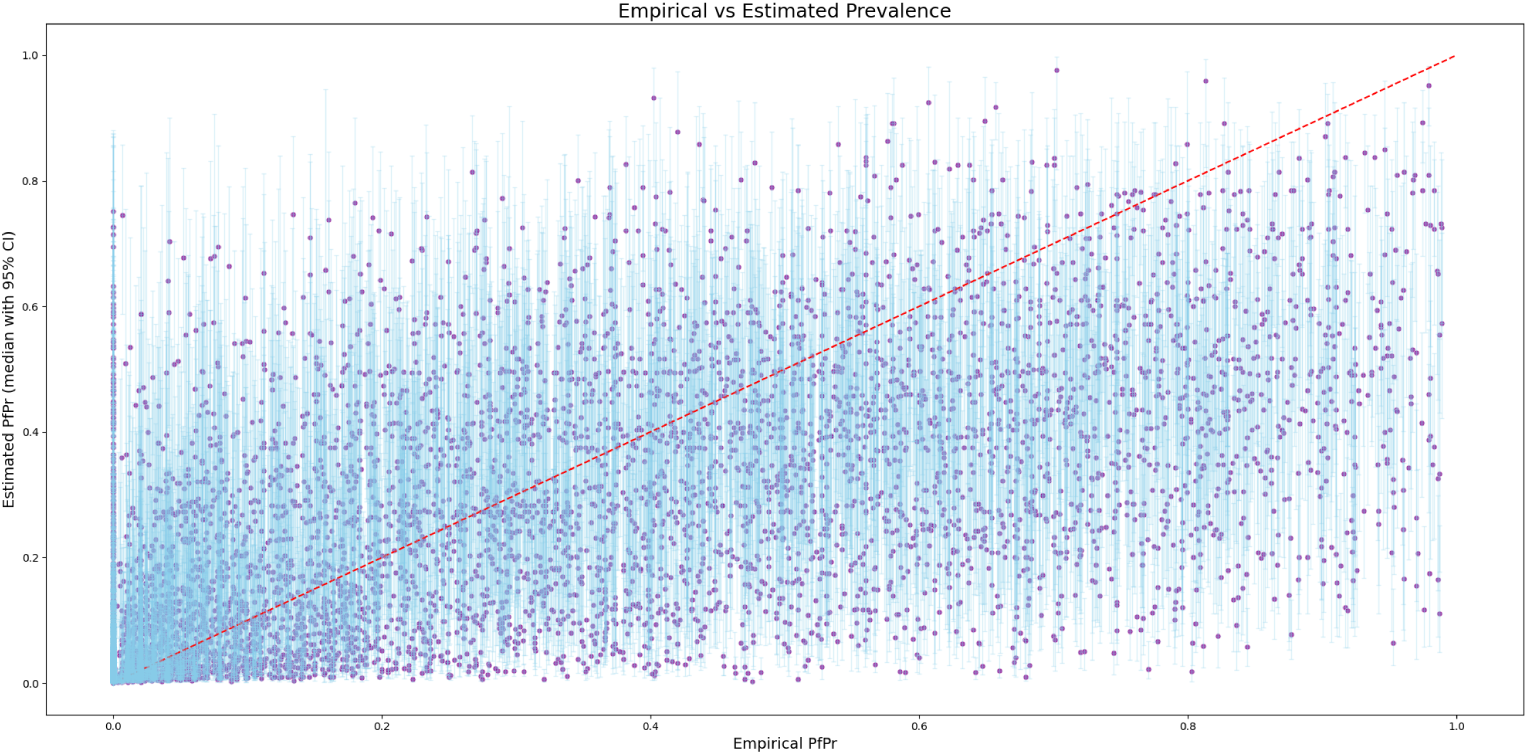
Estimated vs Empirical *PfPr*

We also estimate the amount of baseline malaria cases in each country on an Admin 1 level. The estimates can be found in a CSV file here.

The code for the contrastive learning model and the Bayesian framework can be found on the GitHub repository here.

## Notes

### Competing Interest Statement

The authors have declared no competing interest.

### Author Declarations

The study used Demographic Health Surveys Data- https://www.dhsprogram.com/Data/, World Malaria Report Data- https://www.who.int/teams/global-malaria-programme/reports/world-malaria-report-2024, and LandSat 8 Satellite data- https://developers.google.com/earth-engine/datasets/catalog/landsat

